# Long-term conditions in minoritised urban populations: Latin American community of London

**DOI:** 10.1101/2024.02.20.24303079

**Authors:** James Scuffell, James Bailey, Hiten Dodhia, Stevo Durbaba, Mark Ashworth

**Author notes:** **Corresponding author:** James Scuffell, Department of Population Health Sciences, School of Life Course and Population Sciences, King’s College London, Guy’s Campus, Addison House, London. SE1 1UL.

## Abstract

**Background:** Minoritised populations in the United Kingdom frequently identify in multiple ethnic groupings and therefore little is known of their health needs. There were 136,062 Latin American people recorded in the 2021 UK Census across six different ethnic groups.

**Aim:** Characterise the incidence of long-term conditions (LTCs) and multiple LTCs (mLTCs) amongst the Latin American community of London.

**Design and setting:** Retrospective cohort study using pseudonymised primary care data from 890,922 individuals in an urban, superdiverse area of London from 2005-2022.

**Method:** Latin American individuals were identified using country of birth, language and ethnicity codes, and validated against Census findings. Multivariable competing risks regression models estimated the effect of being Latin American, compared to the White British ethnic group, on incidence of 32 LTCs and risk factors relevant to urban populations.

**Results:** 28,617 Latin American people were identified in this cohort, 3.2% of total. In multivariable analysis, compared to the White British ethnic group, being Latin American was associated with twice the rate of HIV/AIDS (hazard ratio (HR) 2.00; 95% confidence interval (CI) 1.65–2.43), 60% increased rate of diabetes (HR 1.61; 95%CI 1.47–1.77) and almost twice the rate of systemic lupus erythematosus and rheumatoid arthritis (HRs 2.28; 95% CI 1.18–4.38 and 1.69; 95% CI 1.32–2.17 respectively).

**Conclusion:** Using commonly-recorded primary care codes accurately and reliably identifies markedly higher risks of HIV/AIDS, diabetes and joint disease among London’s Latin American population. These data can be used to target inclusive and equitable health interventions.

**How it fits in:** Little is known of the health of the United Kingdom’s (UK) Latin American population, who usually identify in multiple ethnic groups and are therefore hidden from ethnicity statistics. This study identified 28,617 Latin Americans using country of birth, ethnicity and language codes in a large primary care record dataset in London (n=890,438). Compared to the White ethnic group, the Latin American population are at higher risk of HIV/AIDS, diabetes, systemic lupus erythematosus and rheumatoid arthritis. These data reveal health inequalities in a frequently hidden and locally important population of the UK.

## Introduction

The United Kingdom (UK) Census names eighteen ethnic groups in five higher levels: Asian, Black, Mixed, Other and White.^1^ However in 2021 7.5% of those of White ethnicity, and over a quarter of Mixed and Multiple ethnicity respondents, identified in ‘other’ ethnic groups that did not resolve to these eighteen categories.^2^

Minoritised populations in the UK are less likely to access preventative care and they have lower confidence in managing health conditions. Compared to the White British group, Other ethnic groups are less likely to have a recent recorded blood pressure^3^ or HbA1c test.^4^ They are also less likely to be tested for COVID-19^4^ or take up NHS Health Checks.^5^ National data also demonstrate people of Other ethnicities report much lower confidence with self-care and feel less supported in managing health conditions.^6^ They are also substantially less able to mitigate the financial effects of morbidity through sick pay.^7^

### Latin American population of the UK

Stratifying minoritised people into more specific and locally-important categories is important to reveal differential effects of race and ethnicity on health.^8–10^ The Latin American community self-identify as having Latin American origin, with a shared culture and language.^11,12^ By aggregating data from the 288 ‘write-in’ ethnic groups of the 2021 Census, 136,062 UK residents identified as having Latin American ethnicity across the White, Other and Mixed higher level groupings.^13^ In some areas of London they represent 10% of the population.^14^

There is limited research assessing this community’s health needs. 14% of new HIV diagnoses in 2018 were amongst gay and bisexual men (GBM) born in Latin America or the Caribbean.^15^ A search of MEDLINE for studies of Latin American health in the UK yielded one qualitative study on the acceptability of HIV self-testing in London.^16^ Survey data suggest a high proportion of transnational health seeking and some use of private health services.^17^

To improve understanding of health inequalities in this group we developed and validated a phenotype for Latin American ethnicity in routinely-collected primary care data, comparing their life course incidence of long-term conditions (LTCs) to other ethnic groups.

## Method

### Study design

Retrospective cohort study using Lambeth DataNet, a pseudonymised database derived from the electronic health records of adult patients registered to 40 primary care providers in a superdiverse (45% non-White^13^) area of London between 1^st^ April 2005 and 1^st^ February 2022.

For each LTC of interest, individuals were included in a cohort upon registration at a primary care provider or upon turning 18 years of age. They were followed up until an incident LTC, 1^st^ February 2022 (when data collection ceased), deregistration or death. The unit of follow-up was years of age. Individuals were excluded if they were: not resident in London, over 110 years of age; of unknown sex; with a deregistration date after 1^st^ February 2022; with a previous diagnosis of LTC upon cohort registration. Owing to reduced data resolution, participants with less than one year of follow-up were imputed to have six months of follow-up.

### Exposures

Latin American ethnicity was identified in the record using SNOMED-CT codes representing ethnic group, country of birth and main spoken language: (Figure 1). Codelists are available on Figshare and EMIS searches are available on request.^18^

**Figure 1:**
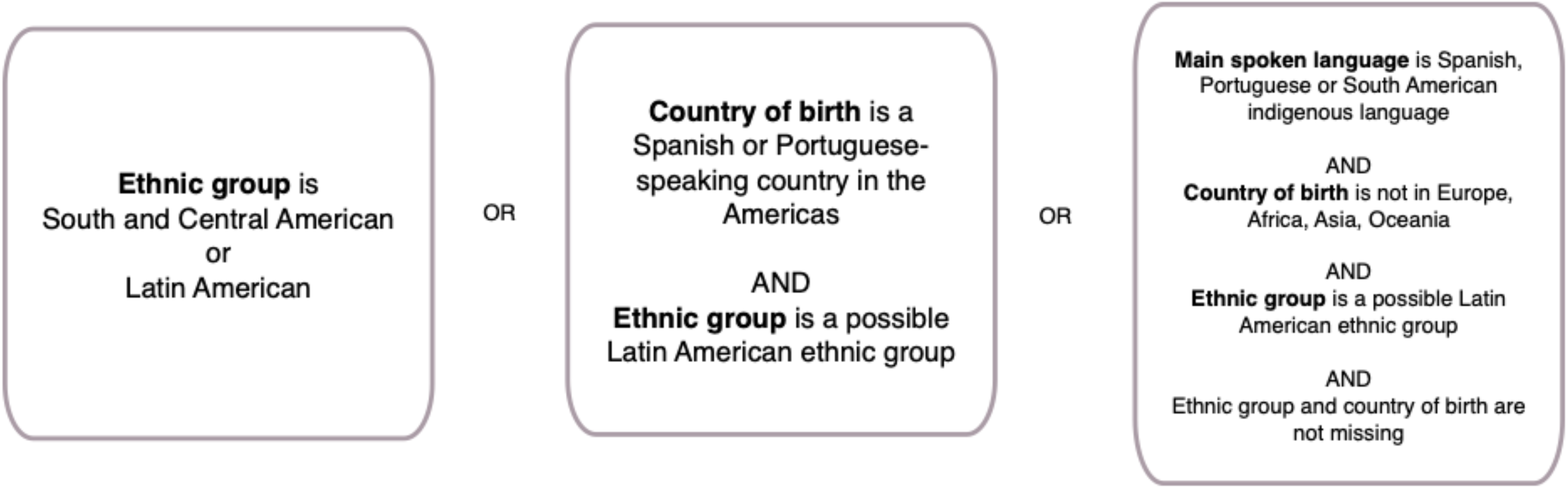
Logic diagram used to phenotype the Latin American population within primary care records.

#### Self-ascribed ethnic group

one author (JS) categorised all ethnic groups into confirmed, possible and unlikely to be assigned to Latin American residents. Where more than one ethnic group was recorded we used the most specific (that is, non-Other) group.

#### Country of birth

majority Spanish- and Portuguese-speaking countries in the Americas.

#### Main spoken language

Spanish; Portuguese; indigenous languages spoken by at least 100,000 inhabitants of Latin America according to United Nations estimates.^19^

We validated this exposure qualitatively with Indoamerican Refugee and Migrant Organisation, a Latin American advocacy charity. We used linear regression and correlation coefficients to determine the extent to which, for each lower super output area (LSOA), the prevalence of Latin American-born people in the 2011 Census^14^ is explained by the proportion of Latin Americans identified in the dataset on 27^th^ March 2011.

We used the aggregated Index of Multiple Deprivation (IMD) 2019 score^20^ for each individual, by either 2011 or 2001 LSOA boundaries. Each LSOA’s IMD score was split into within-borough quintiles.

### Outcomes

We used SNOMED-CT codes for 32 LTCs and 4 risk factors: substance use, obesity, alcohol use (ever having recorded as >14 units/week consumption) and hypercholesterolaemia. These were selected by consensus as locally relevant for ethnically diverse urban populations.^21^ The clinical codes for each LTC are available on request. The first recorded date of each LTC was considered the date of LTC incidence. To reduce the risk of misclassifying prevalent disease as incidence,^22^ we excluded individuals with LTC incidence within 6 months of their cohort start date, or 3 months of registration for those registered on 1^st^ April 2005.

### Statistical analysis

We compared sociodemographic characteristics and LTC cumulative incidence for the Latin American and non-Latin American populations. We calculated the cumulative incidence of multimorbidity (having more than one diagnosed LTC on separate dates) for each group. We used cumulative incidence function plots to visualise the differential acquisition of LTCs in the Latin American, Black and White ethnic groups with increasing age.

For each LTC participants were nested within LSOAs and GP practices. We fitted mixed-effects multivariable Fine-Gray regression models to account for competing risks of death.^23^ We estimated sub-distribution hazard ratios for the Latin American, compared to the White British, ethnic groups, for each LTC. Each model was adjusted *a priori* for age, sex, smoking status (ever/never), and within-borough IMD quintiles. We analysed only patients with a recorded ethnic group, and then conducted sensitivity analyses imputing missing ethnic group data as either White British or Latin American. Analysis was performed in R 4.2.1.

## Results

After data cleaning, a total of 890,438 individuals were included in the dataset for analysis (Figure 2). Participants entered the study at a median 30 years of age (interquartile range (IQR) 25–39), with median follow-up of 4.2 years (IQR 1.8–8.7 years). 25,916 (2.9%) individuals died during follow-up.

**Figure 2:**
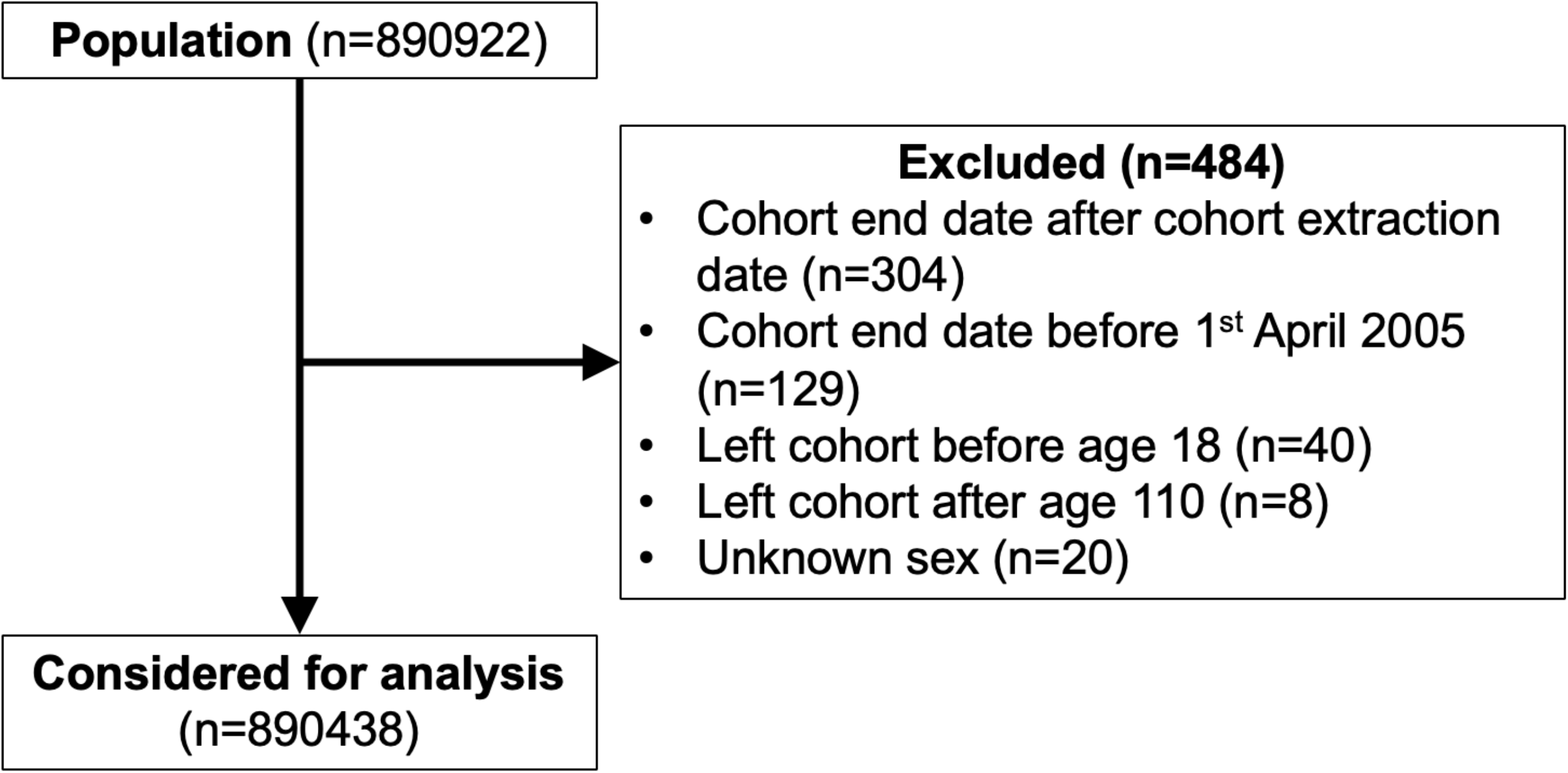
Flowchart demonstrating the total population considered for analysis.

### Missing data

Ethnicity data was missing for 18% (n=164673) of the study population; main spoken language data for 31% (n=274840); country of birth data was missing for 52% (n=464763). 16% had missing data for all three variables; 14% for two variables; 27% for one variable. 386,413 of 890,438 (43%) participants had a recorded ethnic group, main spoken language, and country of birth.

### Latin American population

28,617 Latin American individuals were identified: 3.2% of the cohort. The majority (91%) were identified using country codes, with 2,540 more (9%) identified using ethnic group and 49 by language (Supplementary Data, Figure S1). The Latin American population increased from 3,137 in 2005 to 14,048 in 2022 (Supplementary Data, Figure S2). This is 94% of the 2011 Census-estimated population and 29% more than the 10,833 Latin American-identifying Lambeth residents in the 2021 Census^13^ (Supplementary Data, Figure S3). Most had SNOMED-CT codes representing the UK Census categories of White (45%), Other (37%) or Mixed (13%) (Table S1). Further description of the validation is available in Supplementary Data (Box S1).

Latin American people joined the cohort at a median age of 32.6 versus 30.1 for non-Latin Americans (Table 1). Latin Americans were more likely to reside in deprived LSOAs (27% in the most deprived quintile). 22,185 (85%) recorded a non-English main spoken language, with 57% speaking Spanish and 26% Portuguese (Table 1). Compared to the non-Latin American population, Latin Americans were less likely to have ever smoked, exceeded safe limits of alcohol consumption or to report substance use. (Table 1) Total mortality was 0.9% in the Latin American group versus 3% for non-Latin Americans.

**Table 1:** Sociodemographic characteristics of Latin American and non-Latin American patients, 2005-2022.

|  | Latin American (%) | non-Latin American (%) |
| --- | --- | --- |
| **Total (n=890438)** | 28568 (3.2) | 861870 (96.8) |
| **Age (n=890438)** |  |  |
| 18-30 | 6348 (22.2) | 242230 (28.1) |
| 30-45 | 12614 (44.2) | 374163 (43.4) |
| 45-60 | 7131 (25.0) | 140120 (16.3) |
| 60-80 | 2200 (7.7) | 79954 (9.3) |
| 80+ | 275 (1.0) | 25403 (2.9) |
| **Sex (n=890438)** |  |  |
| Male | 12916 (45.2) | 414289 (48.1) |
| Female | 15652 (54.8) | 447581 (51.9) |
| **Ethnic group (n=725765)** |  |  |
| White | 11848 (44.8) | 472549 (67.6) |
| Black/African/Caribbean/Black British | 1332 (5.0) | 120073 (17.2) |
| Asian/Asian British | 30 (0.1) | 56411 (8.1) |
| Mixed/Multiple ethnic group | 3438 (13.0) | 31598 (4.5) |
| Other ethnic group | 9826 (37.1) | 18660 (2.7) |
| **Deprivation quintile (1=most deprived) (n=890438)** |  |  |
| 1 | 7614 (26.7) | 170474 (19.8) |
| 2 | 7089 (24.8) | 170999 (19.8) |
| 3 | 5499 (19.2) | 172589 (20.0) |
| 4 | 4853 (17.0) | 173234 (20.1) |
| 5 | 3513 (12.3) | 174574 (20.3) |
| **Main spoken language (n=615598)** |  |  |
| English | 3930 (15.0) | 454854 (77.2) |
| Other | 226 (0.9) | 97922 (16.6) |
| Portuguese | 6945 (26.6) | 20443 (3.5) |
| Spanish | 15014 (57.5) | 16264 (2.8) |
| **Ever smoked (n=890438)** |  |  |
| No | 19387 (67.9) | 506464 (58.8) |
| Yes | 9181 (32.1) | 355406 (41.2) |
| **Excess alcohol use (n=890438)** |  |  |
| None | 28251 (98.9) | 831171 (96.4) |
| >14 units/week | 103 (0.4) | 10784 (1.3) |
| Alcohol dependence | 214 (0.7) | 19915 (2.3) |
| **Substance use (n=890438)** |  |  |
| None | 28265 (98.9) | 842727 (97.8) |
| Substance use | 171 (0.6) | 7424 (0.9) |
| Substance dependence | 132 (0.5) | 11719 (1.4) |

### LTCs and mLTCs by ethnic group

In unadjusted analysis, four conditions (systemic lupus erythematosus (SLE), rheumatoid arthritis, HIV/AIDS and hypercholesterolaemia) were more common amongst Latin American people than non-Latin Americans (Figure 3). All other LTCs were less prevalent amongst Latin Americans, with the greatest differences for asthma (4.2% versus 10.6%), anxiety (9.9% versus 15%) and depression (7.1% versus 11.4%). 10.9% of the Latin American group had multimorbidity, compared to 18.0% of non-Latin Americans (Table S5). The predominant LTCs for both groups were chronic pain, anxiety and depression (Figures S5 and S6).

**Figure 3:**
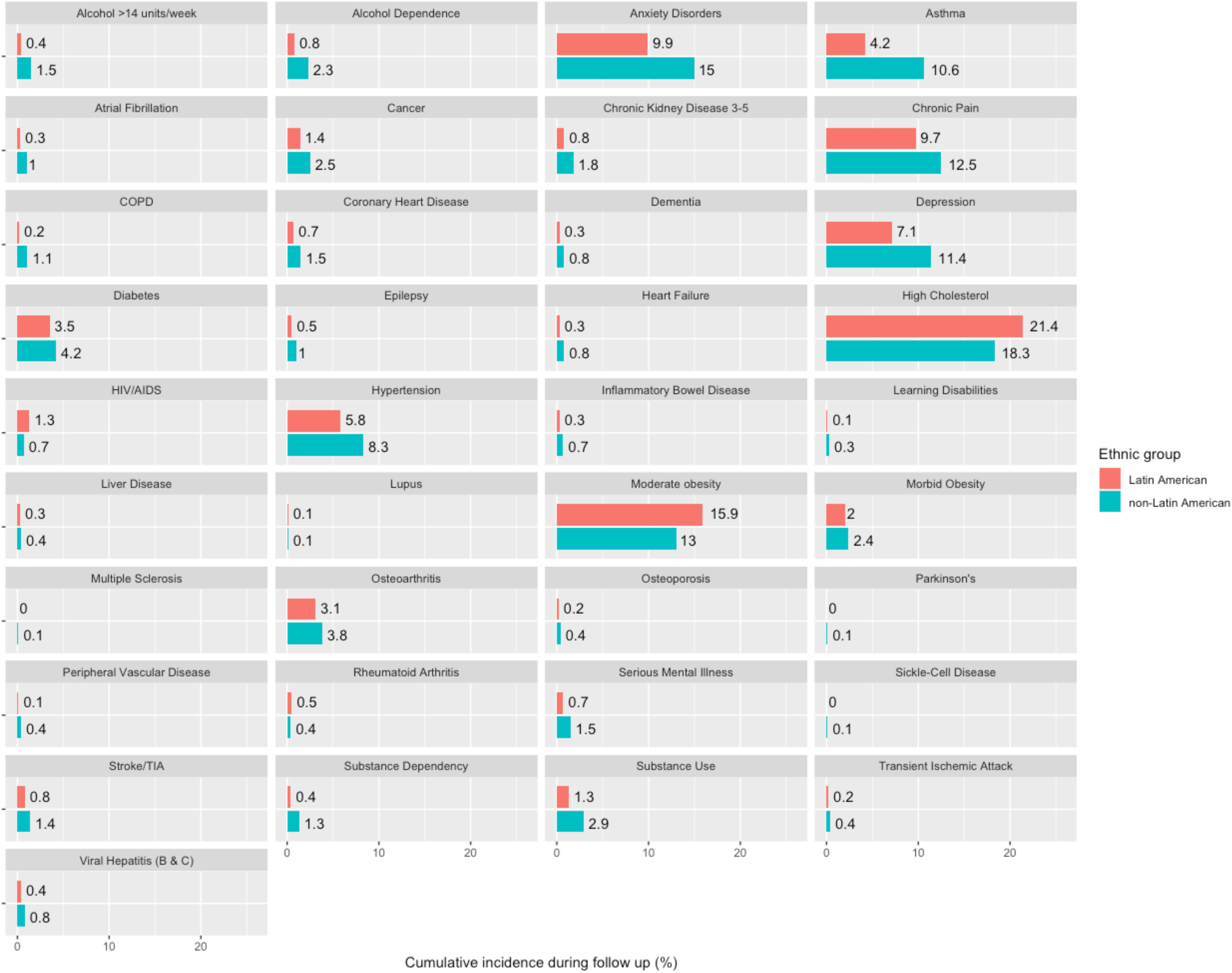
Cumulative incidence of 37 long-term conditions and risk factors during follow up, for Latin American patients and non-Latin Americans. N=890438.

Cumulative incidence plots describe LTC incidence with increasing age for the Black, White and Latin American ethnic groups (n=621190) (Figure 4). Latin Americans have similar rates of cancer, morbid obesity and chronic kidney disease to the White ethnic group. For other conditions – atrial fibrillation, osteoarthritis, inflammatory bowel disease – the risk instead matches the Black group and is lower than the White group. There is an earlier age of incident HIV in the Latin American population compared to Black and White populations.

**Figure 4:**
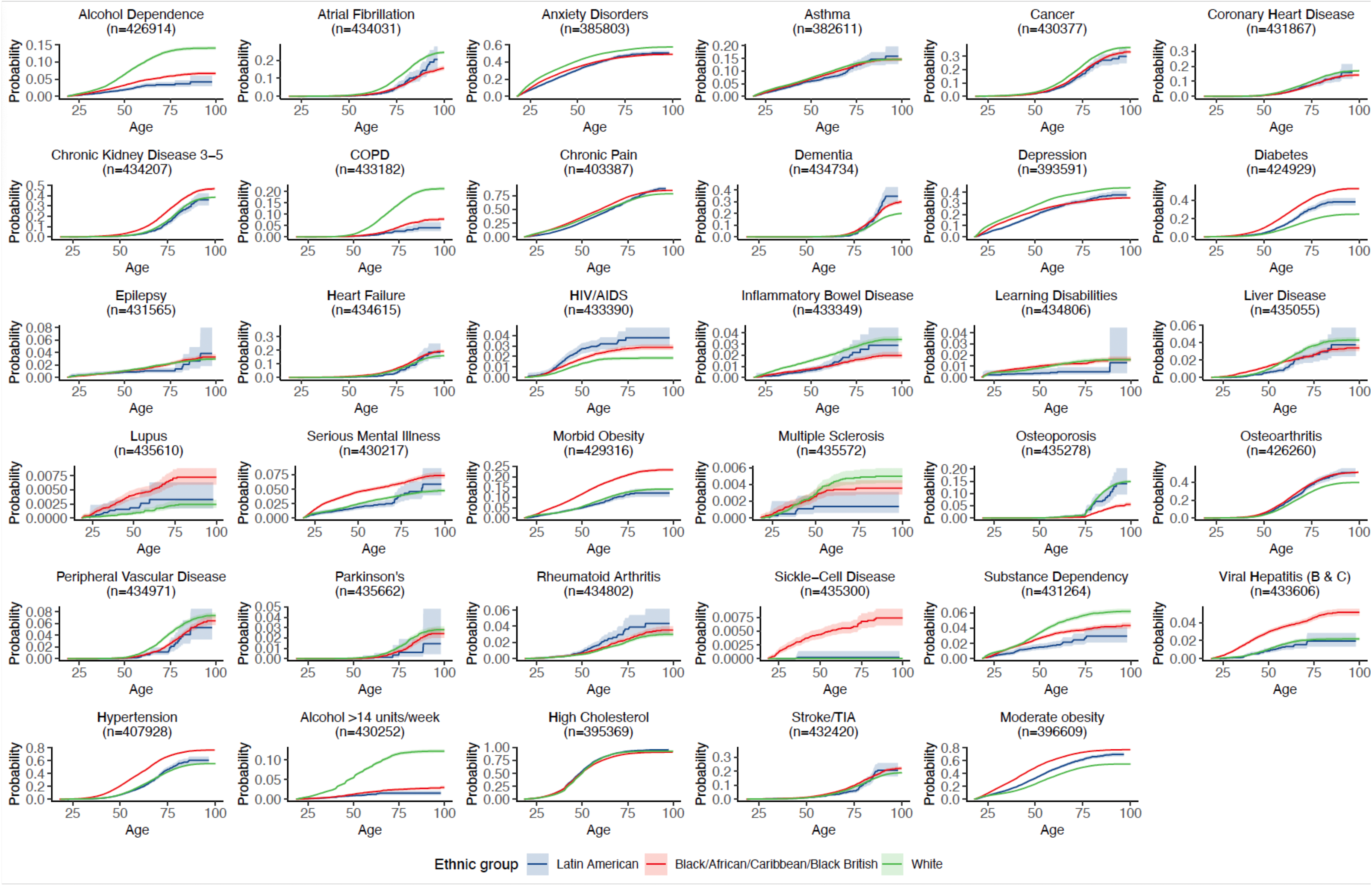
Cumulative incidence function plots for each long-term condition for Latin American-, Black- and White-identifying patients. Shaded areas represent 95% confidence intervals for each curve.

### Multivariable analysis

In multivariable analysis compared to the White British ethnic group (Figure 5, Table S3), HIV/AIDS rates were twice as high among Latin American people (adjusted hazard ratio (HR) 2.00, 95% CI 1.65–2.43).

**Figure 5:**
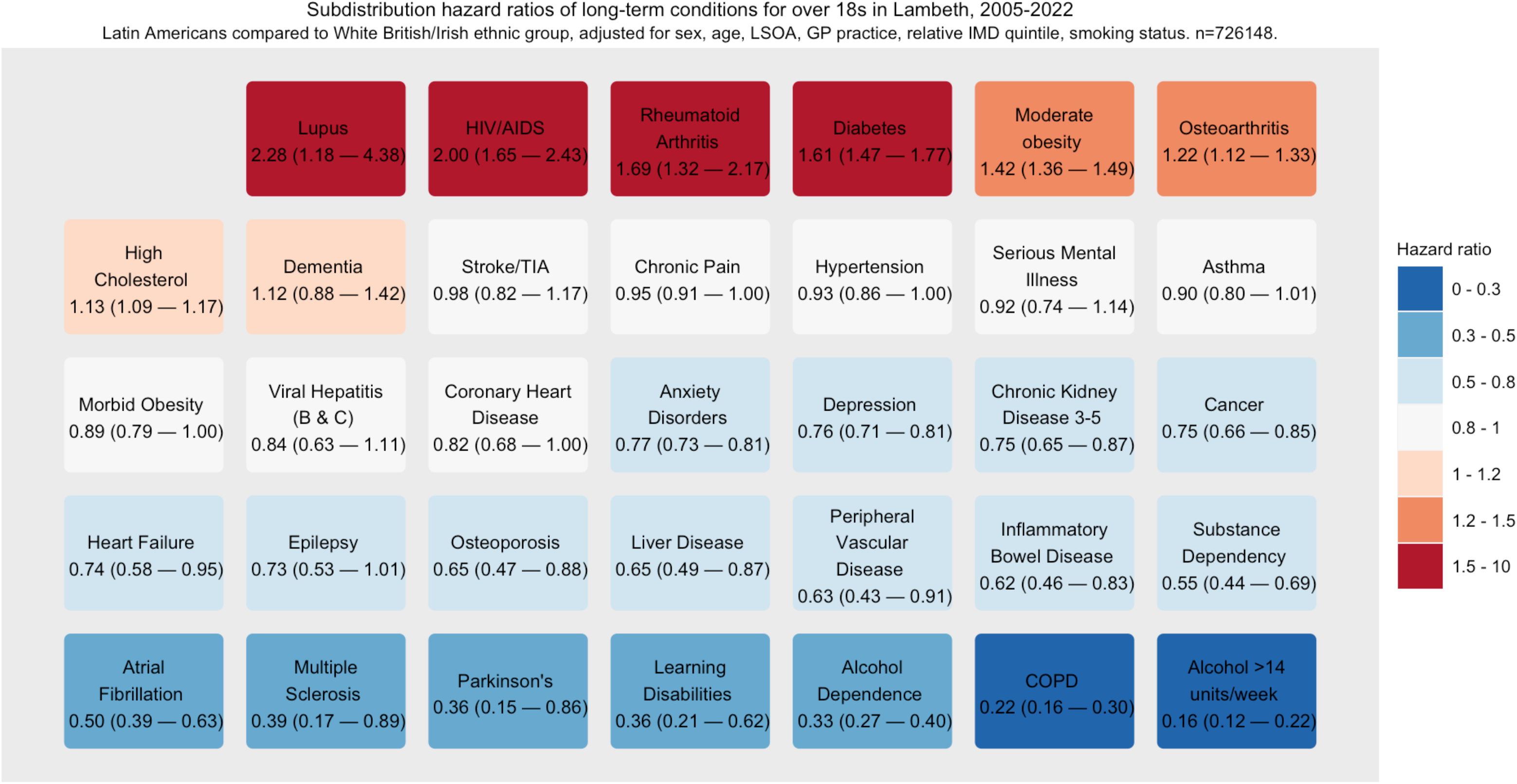
Sub-distribution hazard ratios and 95% confidence intervals for long-term conditions and risk factors for Latin American patients compared to the White ethnic group. Owing to small numbers of sickle cell disease in Latin American patients an estimate was not possible. Competing risks regression adjusted for age, sex, lower super output area, deprivation quintile, smoking status. n=726148.

There was twice the rate of SLE (HR 2.28; 95% CI 1.18–4.38), a 70% increased hazard of rheumatoid arthritis (HR 1.69, 1.32–2.17) and 20% higher recorded osteoarthritis (HR 1.22, 1.12–1.33). There were 60% increased rates of diabetes (HR 1.61, 95% CI 1.47–1.77).

Despite 10% increased rates of hypercholesterolaemia (HR 1.13, 1.09–1.17), there was no evidence of differential rates of stroke or transient ischaemic attack (Figure 5) and there were lower rates of coronary heart disease (HR 0.82, 0.68–1.00). We found statistically strong evidence of lower rates of all cancers (HR 0.75, 0.66–0.85), atrial fibrillation (HR 0.50, 0.39–0.63), and chronic kidney disease stages 3-5 (HR 0.75, 0.65–0.87).

Rates of recorded anxiety and depression were a quarter lower in Latin American people (95% CIs 0.73–0.81 and 0.71–0.81 respectively), with no differential incidence of serious mental illness. Sensitivity analyses (Table S4) demonstrated that the direction of each effect estimate was robust to imputations of ethnic group.

## Discussion

### Summary

This study reveals health inequalities in the Latin American population – a minoritised group who identify across several census ethnic groups. We adjusted for common confounders, providing data to address their disproportionate health challenges.

Latin Americans are at markedly higher risk of developing HIV/AIDS, with higher incidence than the Black ethnic group. This is in the context of Lambeth having the highest HIV prevalence in Europe (1.3%),^24^ and much lower prevalences in most Latin American countries.^25^ As 18% of gay and bisexual men newly diagnosed with HIV in 2018 were Latin American,^26^ this strengthens evidence for HIV transmission occurring in a key population in London.^27^

The Latin American community also has increased obesity, diabetes and hypercholesterolaemia rates, despite low smoking and recorded alcohol consumption. Despite similar rates of hypertension in the White British and Latin American groups, there were lower risks of its cardiovascular sequelae. The markedly lower rates of chronic obstructive pulmonary disease could represent differing occupational exposures or differential measurement of smoking status by general practitioners.^28^

Lower rates of anxiety and depression in young Latin American patients compared to the White and Black ethnic groups may represent underreporting, as self-reported anxiety scores for people of Mixed and Other ethnic groups are consistently higher than for the White ethnic group.^29^ Musculoskeletal conditions are more likely to be diagnosed in the Latin American than White British population.

#### Strengths and limitations

In London one-fifth of Latin American survey respondents have not used primary care,^30^ yet we were able to identify numbers comparable to the 2011 Census and more than the 2021 Census. We identified this many Latin American individuals notwithstanding missing ethnic group information for one fifth of individuals and higher levels of missingness for country of birth and language data. The Latin American phenotype was robust to validation using both country of birth data (2011 Census) and the 288 ‘write-in’ ethnic group categories of the 2021 Census. The dataset is inclusive and representative of the urban population studied, with likely high case ascertainment of LTCs – although this could be improved by using secondary care data. The results were robust to sensitivity analyses. Although results may not be generalisable to rural and less ethnically dense areas, many structural factors affecting Latin American health may apply in other UK urban areas.

Although we adjusted for common biomedical and sociodemographic variables, residual confounding is inevitable, as Latin American people are subject to the unmeasured and intersectional effect of structural, neighbourhood-level, practice-level, and patient-level determinants of health.^31,32^ Language preferences may change over time as migrants become acculturated in the UK; this cannot be accounted for. Finally definitions of race and ethnicity change over time,^8^ and were not longitudinally recorded in these data.

Including other dimensions of ethnicity such as religion and migrant status^9^ might produce a more specific exposure definition. This study was not able to capture individual-level socioeconomic data, which may underestimate neighbourhood and individual determinants of health.^33^

Geographical areas with more Latin American residents tended to have higher-than-expected representation in the dataset, when compared to census data. This was sometimes higher than the average 6% General Practice (GP) over-registration in London.^34^ Further research should identify whether this represents underestimation in the 2021 census, or emigration without deregistration from GP practices. If emigration without deregistration is common this would be an immortal time bias which could underestimate the effect of migrant group status on LTC incidence.^35^

#### Comparison with existing literature

Hispanic Americans have four times the rate of HIV infection, compared to non-Hispanic White Americans.^36^ We found 40% increased rates of diabetes, compared to 65% increased rates of diabetes reported in US Hispanic populations.^37^ The higher rates of lupus and rheumatoid arthritis are also consistent with US data, however given the pathophysiological diversity of SLE^38^ there may be both genetic and structural factors at play.^38–41^ Structural factors affecting the UK Latin American community are likely to be specific to the UK, limiting the ability to compare our findings to those of Hispanic migrants elsewhere.

Low levels of harmful alcohol consumption amongst Latin Americans are also found in US literature, where information on alcohol is not systematically collected.^42^ Although there is no ethnic disparity in recording of alcohol consumption,^43^ it is not clear whether language preference may affect the recording of risk factor information in primary care.

#### Implications for research and practice

This work provides a reproducible methodology to describe chronic health inequalities in minoritised populations over time. Here we identify especially high rates of HIV/AIDS amongst the Latin American community in London, as well as increased rates of rheumatological disorders and diabetes. Further research should include longitudinal multimorbidity clusters^44^ and care quality, from both primary care and secondary care, to better identify health system factors that affect ethnic inequalities in health.

## Data Availability

The data are not publicly available to share, but the research group can provide descriptive aggregate data. Requests should be made to Mark Ashworth.

## Additional information

### Ethical approval

All data were extracted under the terms of a signed data sharing agreement with each practice and with project-specific approval following submission of a data privacy impact assessment, approved by Lambeth Clinical Commissioning Group on 2 November 2017. Information governance approval required ‘low number suppression’. In this case we have suppressed numbers of fewer than 20 patients for ethnic group, and fewer than 100 patients for country of birth tabulations. Separate ethical committee approval was not required (Health Research Authority, 29 September 2017) since all data were fully anonymised for the purposes of research access, and all patient identifiable data had been removed.

## Acknowledgements

The authors would like to acknowledge the contribution of the Indoamerican Refugee and Migrant Organization for their valuable help in validating the codelists used in this study.

## References

1. Ethnic group, national identity and religion - Office for National Statistics. Accessed December 2, 2022. https://www.ons.gov.uk/methodology/classificationsandstandards/measuringequality/ethnicgroupnationalidentityandreligion

2. Ethnic group - Office for National Statistics. Accessed November 30, 2022. https://www.ons.gov.uk/datasets/TS021/editions/2021/versions/1

3. Basta K, Ledwaba-Chapman L, Dodhia H, et al. Hypertension prevalence, coding and control in an urban primary care setting in the UK between 2014 and 2021. Journal of Hypertension.:10.1097/HJH.0000000000003584. doi:10.1097/HJH.0000000000003584

4. Mathur R, Rentsch CT, Morton CE, et al. Ethnic differences in SARS-CoV-2 infection and COVID-19-related hospitalisation, intensive care unit admission, and death in 17 million adults in England: an observational cohort study using the OpenSAFELY platform. The Lancet. 2021;397(10286):1711–1724. doi:10.1016/S0140-6736(21)00634-6

5. Molokhia M, Ayis DS, Karamanos A, et al. What factors influence differential uptake of NHS Health Checks, diabetes and hypertension reviews among women in ethnically diverse South London? Cross-sectional analysis of 63,000 primary care records. EClinicalMedicine. 2022;49:101471. doi:10.1016/j.eclinm.2022.101471

6. Watkinson RE, Sutton M, Turner AJ. Ethnic inequalities in health-related quality of life among older adults in England: secondary analysis of a national cross-sectional survey. The Lancet Public Health. 2021;6(3):e145–e154. doi:10.1016/S2468-2667(20)30287-5

7. Patel P, Beale S, Nguyen V, et al. Inequalities in access to paid sick leave among workers in England and Wales. Int J Health Plann Manage. 2023;38(6):1864–1876. doi:10.1002/hpm.3697

8. Selvarajah S, Deivanayagam TA, Lasco G, et al. Categorisation and Minoritisation. BMJ Glob Health. 2020;5(12):e004508. doi:10.1136/bmjgh-2020-004508

9. Aspinall PJ. The utility and validity for public health of ethnicity categorization in the 1991, 2001 and 2011 British Censuses. Public Health. 2011;125(10):680–687. doi:10.1016/j.puhe.2011.05.001

10. Aspinall PJ. Operationalising the collection of ethnicity data in studies of the sociology of health and illness. Sociology of Health & Illness. 2001;23(6):829–862. doi:10.1111/1467-9566.00277

11. Bhopal R. Glossary of terms relating to ethnicity and race: for reflection and debate. Journal of Epidemiology & Community Health. 2004;58(6):441–445. doi:10.1136/jech.2003.013466

12. Real Academia Española. Diccionario de la lengua española. 23.6.; 2022. Accessed January 5, 2023. https://dle.rae.es/latinoamericano?m=form

13. Ethnic group (detailed) - Office for National Statistics. Accessed November 29, 2022. https://www.ons.gov.uk/datasets/TS022/editions/2021/versions/1?showAll=ethnic_group_288a#get-data

14. QS203EW (Country of birth (detailed)) - Nomis - Official Labour Market Statistics. Accessed April 6, 2022. https://www.nomisweb.co.uk/census/2011/qs203ew

15. O’Halloran C, Sun S, Nash S, et al. HIV in the United Kingdom: Towards Zero 2030. 2019 Report. Public Health England; 2019. Accessed January 8, 2023. https://assets.publishing.service.gov.uk/government/uploads/system/uploads/attachment_data/file/965765/HIV_in_the_UK_2019_towards_zero_HIV_transmissions_by_2030.pdf

16. Nicholls EJ, Samba P, McCabe L, et al. Experiences of and attitudes towards HIV testing for Asian, Black and Latin American men who have sex with men (MSM) in the SELPHI (HIV Self-Testing Public Health Intervention) randomized controlled trial in England and Wales: implications for HIV self-testing. BMC Public Health. 2022;22(1):809. doi:10.1186/s12889-022-13189-7

17. Gideon J. Exploring migrants’ health seeking strategies: the case of Latin American migrants in London. International Journal of Migration, Health and Social Care. 2011;7(4):197–208. doi:10.1108/17479891111206328

18. Scuffell et al. Latin American Codelists. Published online April 6, 2023. doi:10.6084/m9.figshare.22438855.v1

19. United Nations Statistics Division. Population by language, sex and urban/rural residence. Published 2022. Accessed April 6, 2022. http://data.un.org/Data.aspx?d=POP&f=tableCode:27

20. MHCLG Open Data : English Indices of Deprivation 2019 - LSOA Level. Accessed April 14, 2022. https://opendatacommunities.org/data/societal-wellbeing/imd2019/indices

21. Hafezparast N, Turner EB, Dunbar-Rees R, et al. Adapting the definition of multimorbidity – development of a locality-based consensus for selecting included Long Term Conditions. BMC Family Practice. 2021;22(1):124. doi:10.1186/s12875-021-01477-x

22. Lewis JD, Bilker WB, Weinstein RB, Strom BL. The relationship between time since registration and measured incidence rates in the General Practice Research Database. Pharmacoepidemiology and Drug Safety. 2005;14(7):443–451. doi:10.1002/pds.1115

23. Fine JP, Gray RJ. A Proportional Hazards Model for the Subdistribution of a Competing Risk. Journal of the American Statistical Association. 1999;94(446):496–509. doi:10.1080/01621459.1999.10474144

24. Annex B: Local authorities with high or very high HIV prevalence (2019). GOV.UK. Accessed November 4, 2022. https://www.gov.uk/government/publications/towards-zero-the-hiv-action-plan-for-england-2022-to-2025/annex-b-local-authorities-with-high-or-very-high-hiv-prevalence-2019

25. Joint United Nations Programme on HIV/AIDS. UNAIDS Data 2023. Joint United Nations Programme on HIV/AIDS; 2023. Accessed December 18, 2023. https://www.unaids.org/en/resources/documents/2023/2023_unaids_data

26. Rawson S, Croxford S, Swift B, Kirwan P, Guerra L, Delpech V. Latin Americans in the UK: a key population for HIV prevention. In: HIV Medicine. Vol 20 Suppl 5.; 2019:16–75. Accessed January 8, 2023. https://www.bhiva.org/file/5ca469a56b895/AbstractBook2019.pdf

27. Public Health England. Trends in HIV Testing, New Diagnoses and People Receiving HIV-Related Care in the United Kingdom: Data to the End of December 2019. Public Health England; 2020. Accessed December 9, 2022. https://assets.publishing.service.gov.uk/government/uploads/system/uploads/attachment_data/file/959330/hpr2020_hiv19.pdf

28. Atkinson MD, Kennedy JI, John A, Lewis KE, Lyons RA, Brophy ST. Development of an algorithm for determining smoking status and behaviour over the life course from UK electronic primary care records. BMC Med Inform Decis Mak. 2017;17:2. doi:10.1186/s12911-016-0400-6

29. Office of National Statistics. Anxiety by ethnicity. Published 2019. Accessed December 15, 2022. https://www.ethnicity-facts-figures.service.gov.uk/health/wellbeing/well-being-anxiety-yesterday/latest

30. McIllwaine C, Camilo Cock J, Linneker B. No Longer Invisible: The Latin American Community in London. Queen Mary, University of London; 2011. Accessed December 15, 2022. https://trustforlondon.fra1.cdn.digitaloceanspaces.com/media/documents/No-Longer-Invisible-report.pdf

31. Selvarajah S, Maioli SC, Deivanayagam TA, et al. Racism, xenophobia, and discrimination: mapping pathways to health outcomes. The Lancet. 2022;400(10368):2109–2124. doi:10.1016/S0140-6736(22)02484-9

32. Kaplan JB, Bennett T. Use of race and ethnicity in biomedical publication. JAMA. 2003;289(20):2709–2716. doi:10.1001/jama.289.20.2709

33. Nazroo JY. The Structuring of Ethnic Inequalities in Health: Economic Position, Racial Discrimination, and Racism. Am J Public Health. 2003;93(2):277–284.

34. Burch P, Doran T, Kontopantelis E. Regional variation and predictors of over-registration in English primary care in 2014: a spatial analysis. J Epidemiol Community Health. 2018;72(6):532–538. doi:10.1136/jech-2017-210176

35. Lévesque LE, Hanley JA, Kezouh A, Suissa S. Problem of immortal time bias in cohort studies: example using statins for preventing progression of diabetes. BMJ. 2010;340:b5087. doi:10.1136/bmj.b5087

36. Centers for Disease Control and Prevention (CDC). HIV Surveillance Report 2019.; 2021. Accessed December 18, 2023. https://www.cdc.gov/hiv/pdf/library/reports/surveillance/cdc-hiv-surveillance-report-2018-updated-vol-32.pdf

37. Centers for Disease Control and Prevention (CDC). Summary Health Statistics: National Health Interview Survey, 2018. Table A-4a. Published online 2018. Accessed December 2, 2022. https://ftp.cdc.gov/pub/Health_Statistics/NCHS/NHIS/SHS/2018_SHS_Table_A-4.pdf

38. Lewis MJ, Jawad AS. The effect of ethnicity and genetic ancestry on the epidemiology, clinical features and outcome of systemic lupus erythematosus. Rheumatology. 2017;56(suppl_1):i67–i77. doi:10.1093/rheumatology/kew399

39. Grubbs V, Cerdeña JP, Non AL. The misuse of race in the search for disease-causing alleles. The Lancet. 2022;399(10330):1110–1111. doi:10.1016/S0140-6736(22)00488-3

40. Xu Y, Wu Q. Prevalence Trend and Disparities in Rheumatoid Arthritis among US Adults, 2005–2018. J Clin Med. 2021;10(15):3289. doi:10.3390/jcm10153289

41. Molina E, Haas R, del Rincon I, Battafarano DF, Restrepo JF, Escalante A. Does the “Hispanic Paradox” apply to Rheumatoid Arthritis? Survival Data from a Multi-Ethnic Cohort. Arthritis Care Res (Hoboken). 2014;66(7):972–979. doi:10.1002/acr.22254

42. Castañeda SF, Garcia ML, Lopez-Gurrola M, et al. Alcohol use, acculturation and socioeconomic status among Hispanic/Latino men and women: The Hispanic Community Health Study/Study of Latinos. PLOS ONE. 2019;14(4):e0214906. doi:10.1371/journal.pone.0214906

43. Mansfield K, Crellin E, Denholm R, et al. Completeness and validity of alcohol recording in general practice within the UK: a cross-sectional study. BMJ Open. 2019;9(11):e031537. doi:10.1136/bmjopen-2019-031537

44. Soley-Bori M, Bisquera A, Ashworth M, et al. Identifying multimorbidity clusters with the highest primary care use: 15 years of evidence from a multi-ethnic metropolitan population. Br J Gen Pract. 2022;72(716):e190–e198. doi:10.3399/BJGP.2021.0325

