## Supplementary data for "Long-term conditions in minoritised urban populations: Latin American community of London"

### Box S1: Validating the Latin American phenotype

There were 8,335 Latin American Lambeth residents in the cohort on 27th March 2011 – 3.2% of the total. The dataset-registered population grew to 14,048 at the beginning of 2022 (Figure S2). This compared to 9,352 Latin American-born residents identified in Lambeth in the 2011 census (3.1%) and 9,678 Latin American-identifying residents in the 2021 census (3.1%)[1]. When the Latin American population count was measured at lower super-output area there was a strong correlation between datasets, with the cohort ascertaining 94% of the census-estimated Latin American-born population (Figure S3, regression coefficient 0.94, $R^2$=0.71). Areas in which there was a relative over-registration of Latin Americans in the cohort tended to be in areas with higher Latin American-born populations. LSOAs with relative under-registration of Latin Americans tended to be close the boundaries of the borough, and with lower Latin American-born populations (Figure S4).

We worked with the Indoamerican Refugee and Migrant Organization to qualitatively assess concordance between self-ascribed Latin American ethnicity and the codes used to identify Latin American patients in the health record.

1 Ethnic group (detailed) - Office for National Statistics. https://www.ons.gov.uk/datasets/TS022/editions/2021/versions/1?showAll=ethnic_group_288a#get-data (accessed 29 Nov 2022).

Figure S1: Venn diagram showing the types of codes used to identify Latin American people within the dataset

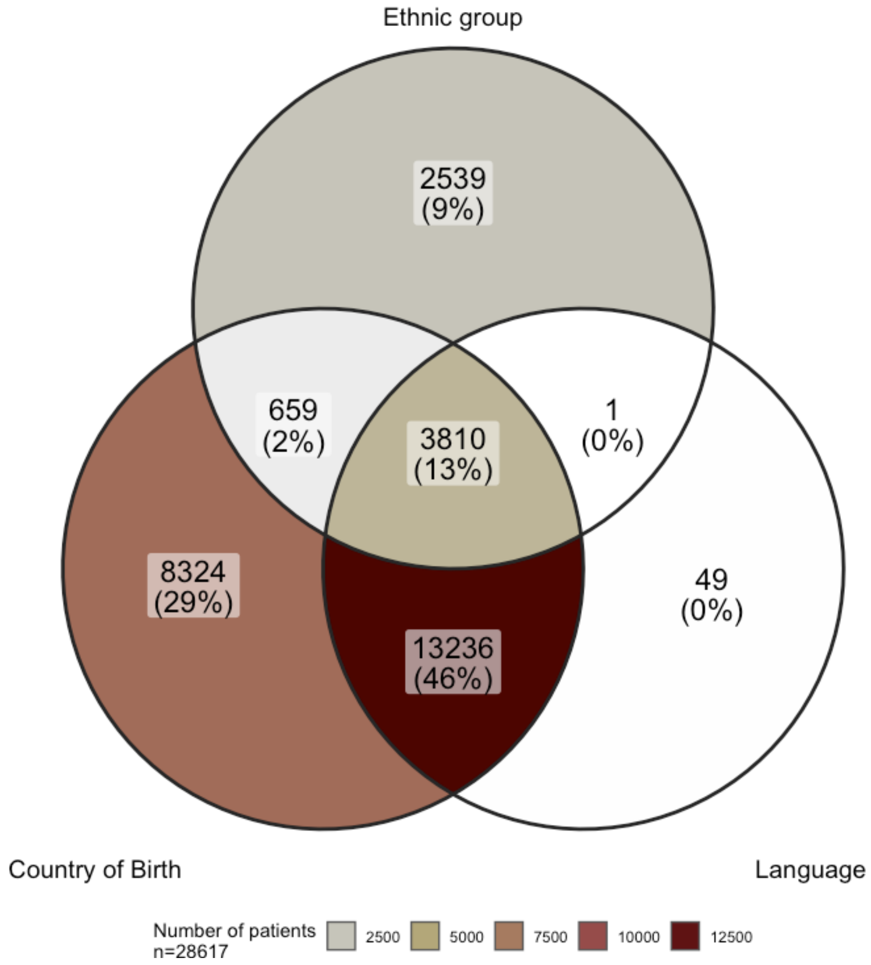

Figure S2: Number of Latin American patients in Lambeth DataNet, 2005-2022.

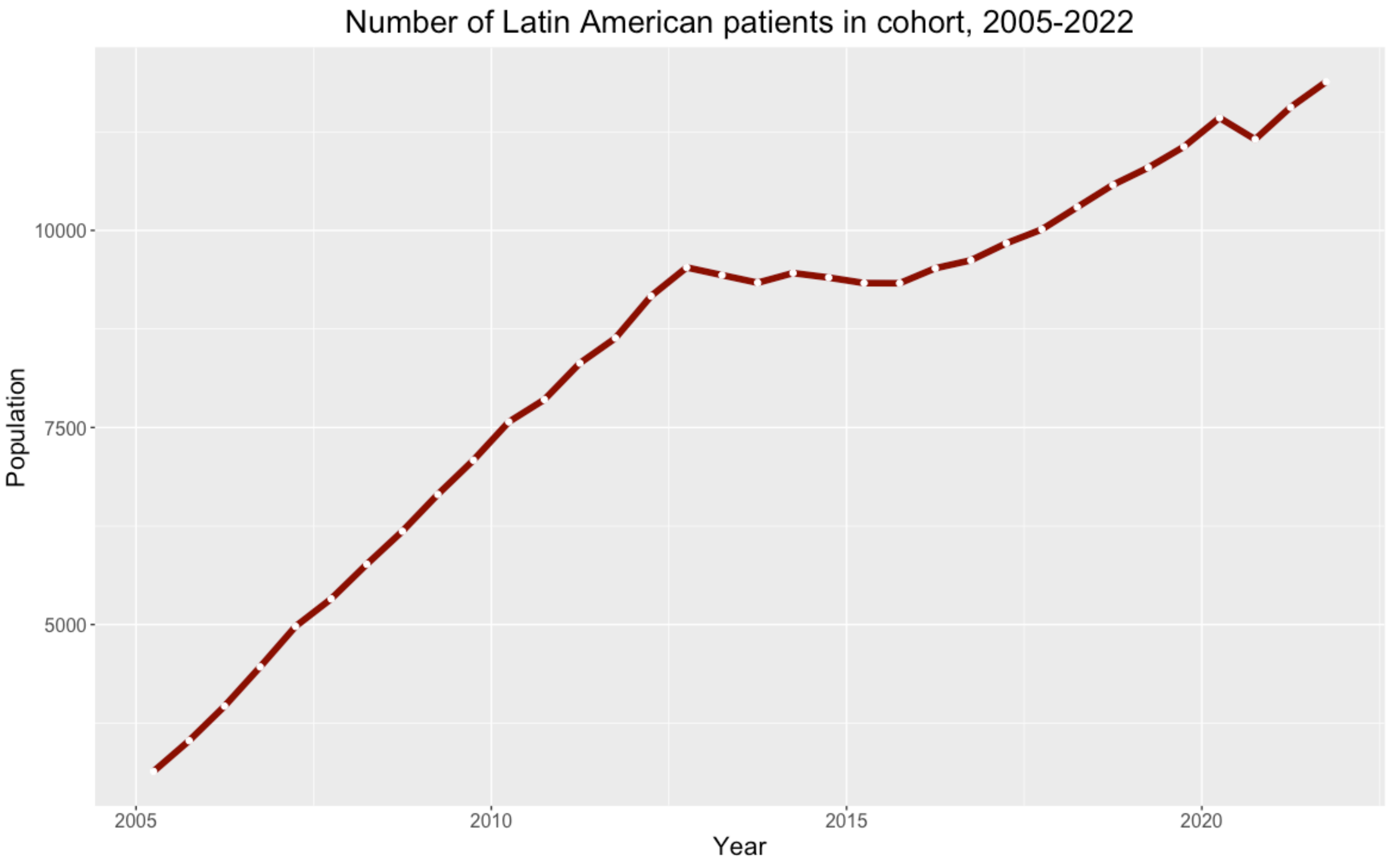

Figure S3: Number of Latin American-born residents of Lambeth on census day 2011. UK Census data, compared to Lambeth DataNet cohort. Each point represents a lower super-output area (LSOA).

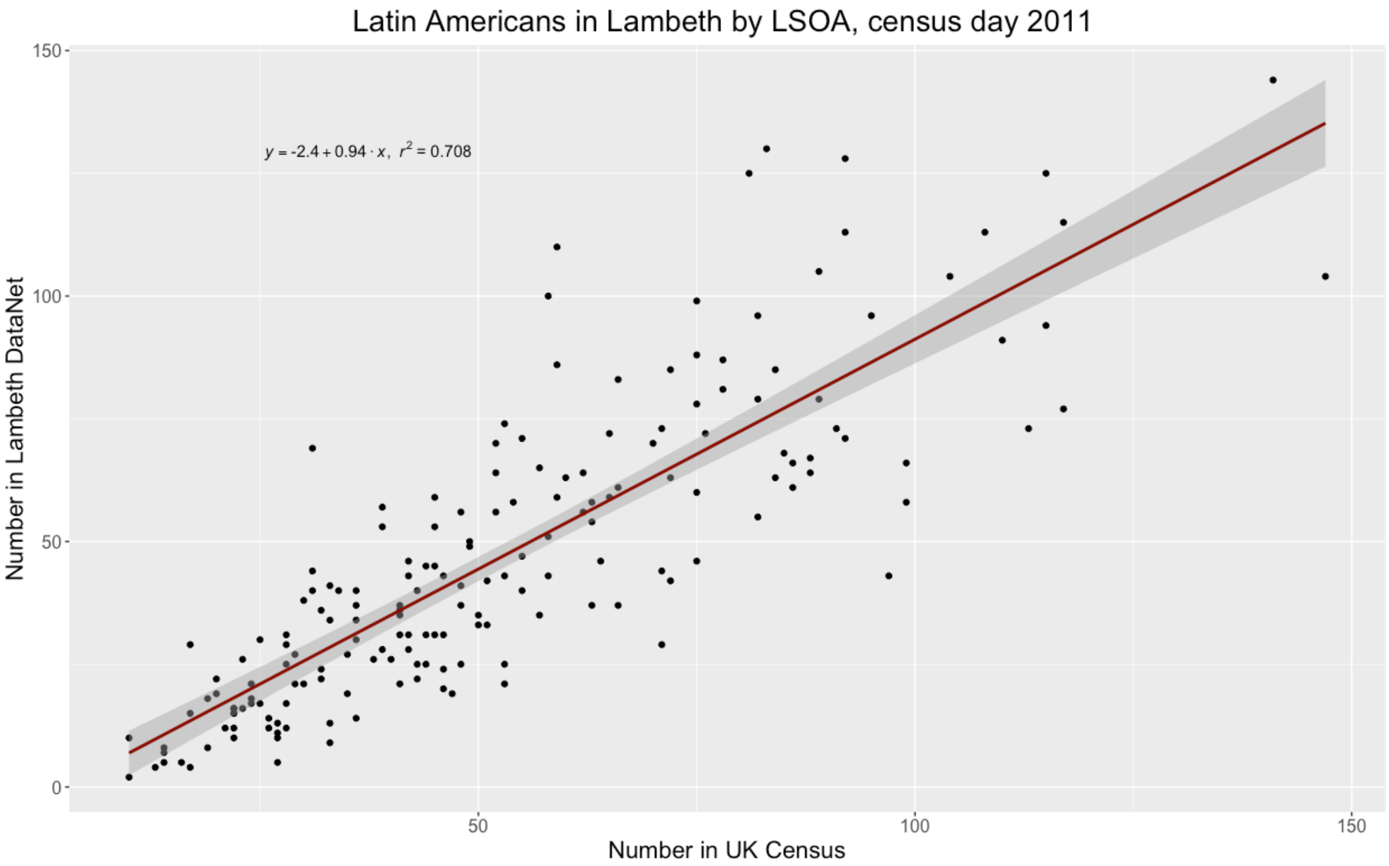

Figure S4: Choropleth map showing areas of GP over-registration and under-registration of Latin American patients in Lambeth, compared to 2011 census data. Map tiles are © OpenStreetMap contributors and used under a Creative Commons Attribution-ShareAlike 2.0 license.

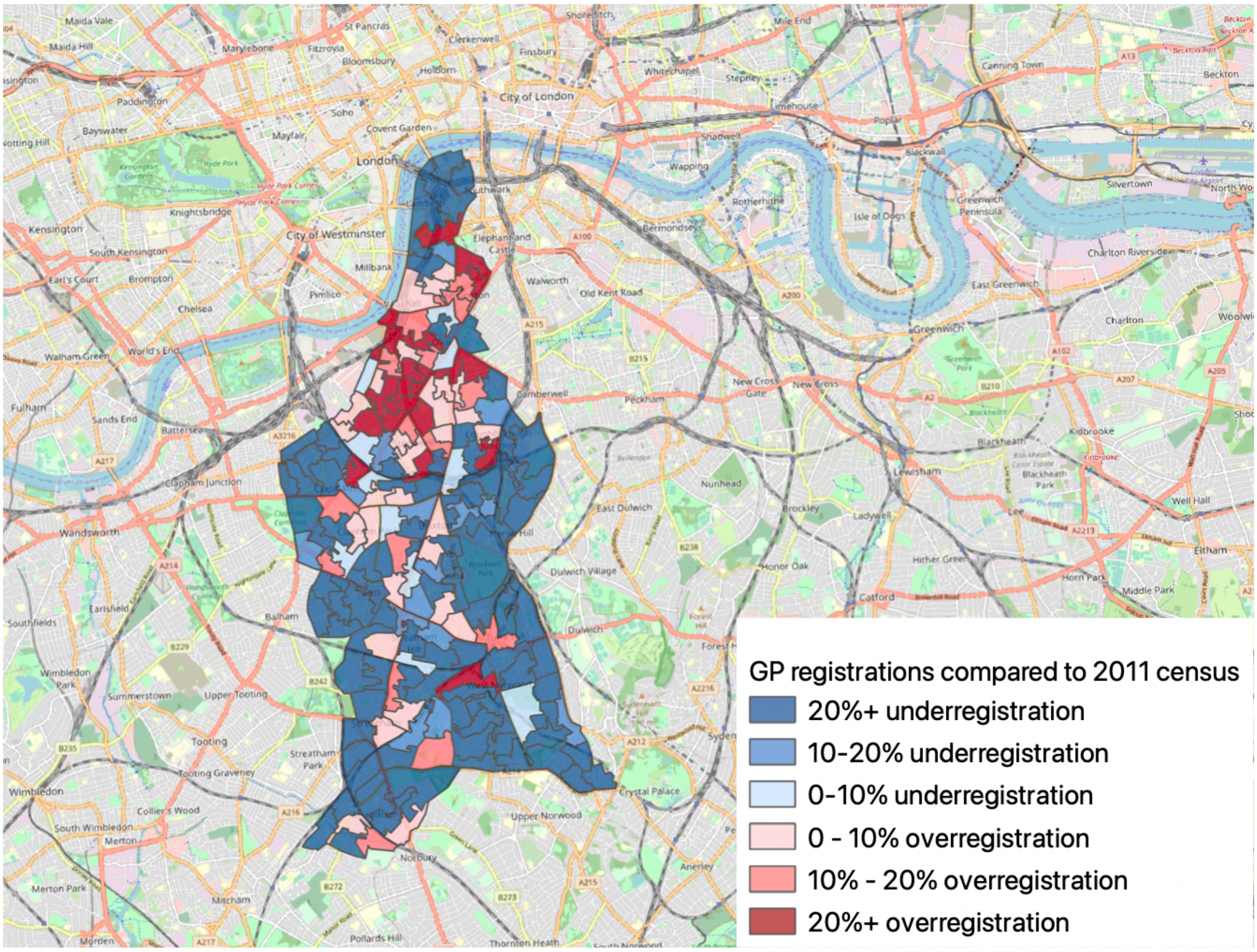

Figure S5: UpSet plot showing the most common thirty long-term conditions (LTCs) and clusters of LTCs recorded in the Latin American and non-Latin American populations, 2005-2022.

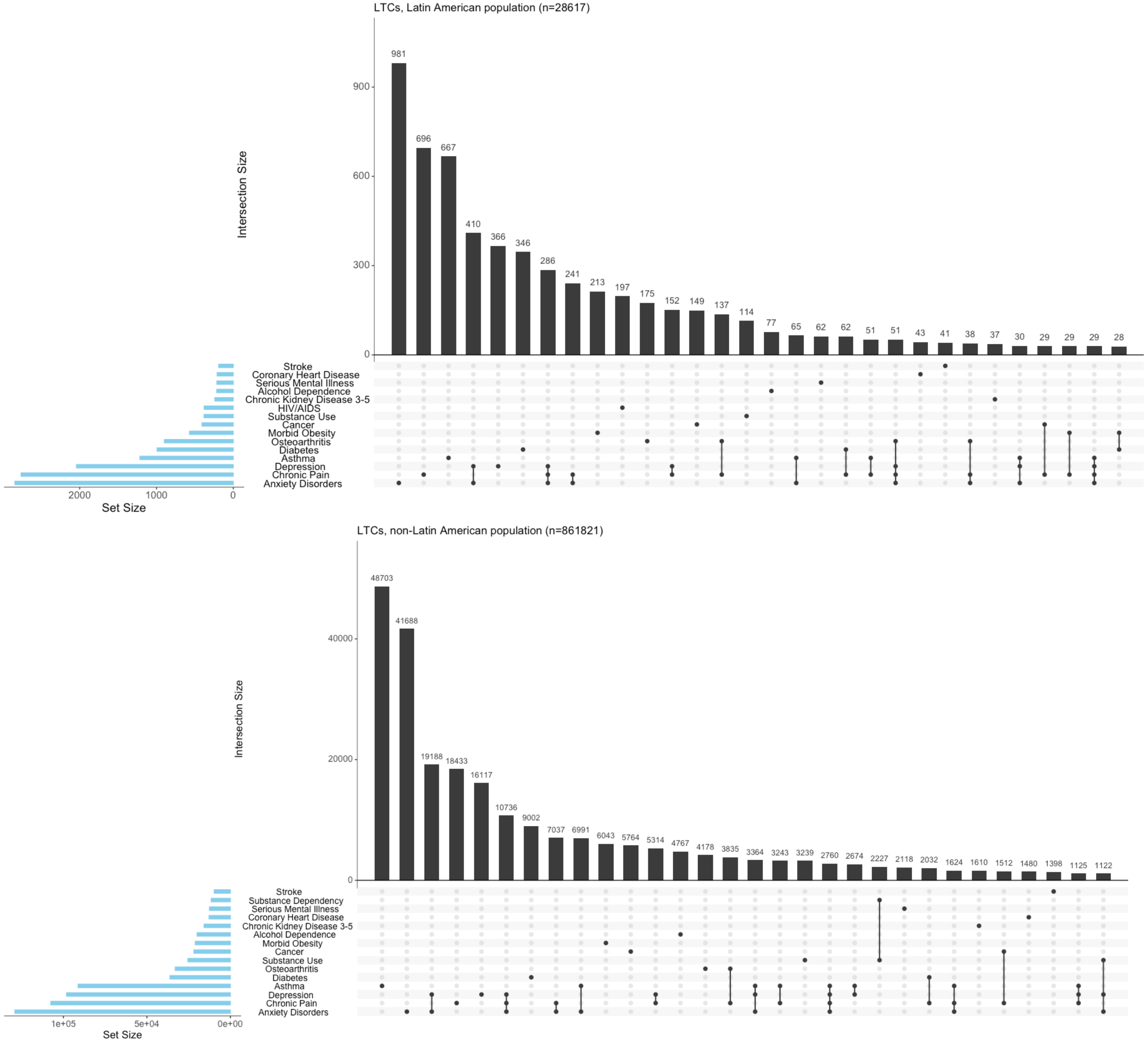

Figure S6: UpSet plot showing the most commonly recorded multiple long term condition clusters in the Latin American and non-Latin American populations, 2005-2022.

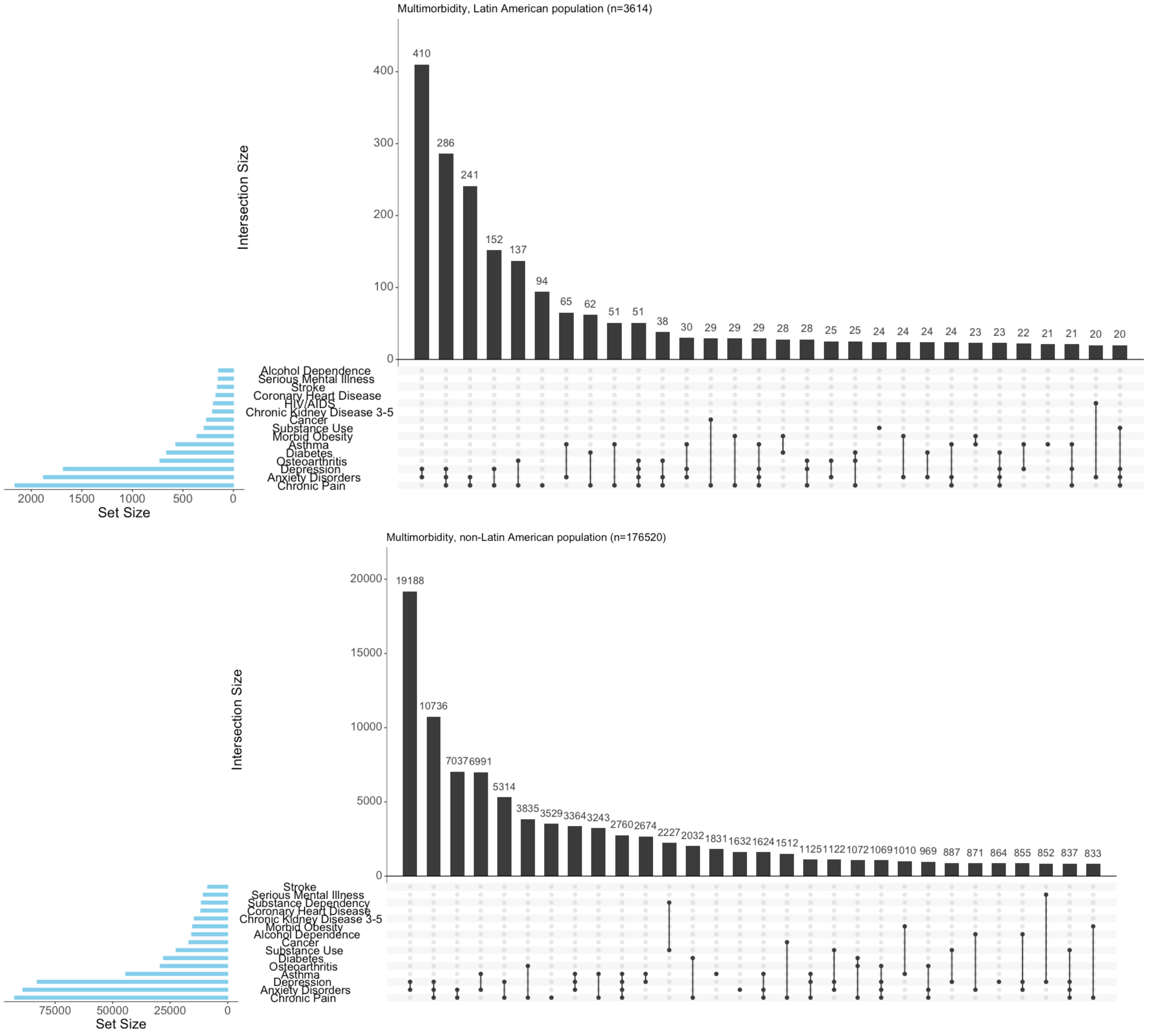

Table S1: Proportion of Latin American individuals with SNOMED-CT codes for UK Census ethnic groups. Only ethnic groups containing more than 20 people are included.

| **Ethnic group** | **n (% of total)** | **Ethnic group** | **n (% of total)** |
|---|---|---|---|
| White | 11874 (41.5) | Other White | 11483 (40.1) |
| | | British | 389 (1.4) |
| Other ethnic group | 9832 (34.4) | Any other ethnic group | 9832 (34.4) |
| Mixed/Multiple ethnic group | 3449 (12.1) | Other Mixed | 2824 (9.9) |
| | | White and Black Caribbean | 602 (2.1) |
| Black | 1338 (4.7) | Caribbean | 997 (3.5) |
| | | Other Black | 326 (1.1) |
| Asian/Asian British | 30 (0.1) | | |
| *Missing* | *2094 (7.3)* | | |
| **Total** | **28617 (100%)** | | |

Table S2: Proportion of Latin American people in dataset by SNOMED-CT codes for country of birth.

| Country of birth | n (% of total) |
|---|---|
| Brazil | 8524 (29.8) |
| Columbia | 5795 (20.3) |
| Ecuador | 4277 (14.9) |
| Missing | 2589 (9) |
| Bolivia | 1502 (5.2) |
| Dominican Republic | 1055 (3.7) |
| Peru | 889 (3.1) |
| Venezuela | 888 (3.1) |
| Argentina | 758 (2.6) |
| Mexico | 572 (2) |
| Guyana | 560 (2) |
| Chile | 391 (1.4) |
| Honduras | 173 (0.6) |
| Cuba | 150 (0.5) |
| Paraguay | <100 (<0.5) |
| Nicaragua | <100 (<0.5) |
| Uruguay | <100 (<0.5) |
| El Salvador | <100 (<0.5) |
| Guatemala | <100 (<0.5) |
| Costa Rica | <100 (<0.5) |
| Panama | <100 (<0.5) |
| Suriname | <100 (<0.5) |
| **Total** | **28617 (100%)** |

Table S3: Multivariable hazard ratios of 36 long-term conditions and risk factors. Each column is derived from a separate mixed effects competing risks regression model, with random effects terms for GP practice and lower super output area. Adjustment is made for sex, smoking and within-dataset deprivation quintile. N=579086.

| Variable | Alcohol Dependence | Atrial Fibrillation | Alcohol >14 units/week | Anxiety Disorders | Asthma | Cancer | Coronary Heart Disease | High Cholesterol | Chronic Kidney Disease 3-5 | COPD | Chronic Pain | Dementia |
|---|---|---|---|---|---|---|---|---|---|---|---|---|
| Total events | 6469 | 5335 | 4841 | 49561 | 9062 | 11703 | 5285 | 88276 | 12309 | 4899 | 55589 | 4456 |
| **Sex** | | | | | | | | | | | | |
| Male | - | - | - | - | - | - | - | - | - | - | - | - |
| Female | 0.53 (0.5 - 0.56) | 0.83 (0.79 - 0.88) | 0.4 (0.37 - 0.42) | 1.96 (1.93 - 2) | 1.48 (1.42 - 1.55) | 0.96 (0.93 - 1) | 0.59 (0.56 - 0.63) | 1.22 (1.2 - 1.23) | 1.32 (1.27 - 1.37) | 0.94 (0.89 - 1) | 1.83 (1.8 - 1.86) | 1.42 (1.33 - 1.51) |
| **Ethnic group** | | | | | | | | | | | | |
| White British | - | - | - | - | - | - | - | - | - | - | - | - |
| Black/African/Caribbean/Black British | 0.52 (0.49 - 0.56) | 0.5 (0.47 - 0.55) | 0.27 (0.24 - 0.29) | 0.83 (0.81 - 0.86) | 0.95 (0.89 - 1) | 0.82 (0.78 - 0.86) | 0.76 (0.71 - 0.82) | 0.92 (0.9 - 0.94) | 1.39 (1.33 - 1.46) | 0.32 (0.3 - 0.35) | 1.13 (1.11 - 1.16) | 1.32 (1.23 - 1.43) |
| Asian/Asian British | 0.37 (0.32 - 0.43) | 0.55 (0.49 - 0.62) | 0.22 (0.18 - 0.26) | 0.76 (0.73 - 0.79) | 1.25 (1.16 - 1.35) | 0.59 (0.54 - 0.64) | 1.72 (1.57 - 1.89) | 1.19 (1.16 - 1.23) | 1.17 (1.09 - 1.26) | 0.51 (0.44 - 0.59) | 1.02 (0.98 - 1.05) | 1 (0.88 - 1.14) |
| Mixed/Multiple ethnic group | 0.74 (0.66 - 0.84) | 0.61 (0.5 - 0.74) | 0.44 (0.37 - 0.53) | 1.01 (0.97 - 1.06) | 1.17 (1.06 - 1.3) | 0.81 (0.72 - 0.9) | 1 (0.85 - 1.18) | 0.97 (0.94 - 1.01) | 1.19 (1.07 - 1.32) | 0.58 (0.48 - 0.69) | 1.1 (1.05 - 1.15) | 1.28 (1.07 - 1.54) |
| Other ethnic group | 0.53 (0.44 - 0.63) | 0.65 (0.51 - 0.83) | 0.24 (0.18 - 0.32) | 0.8 (0.75 - 0.85) | 0.92 (0.8 - 1.06) | 0.77 (0.66 - 0.89) | 1.19 (0.99 - 1.44) | 0.95 (0.9 - 0.99) | 0.91 (0.78 - 1.06) | 0.51 (0.4 - 0.65) | 0.91 (0.86 - 0.96) | 1.01 (0.77 - 1.35) |
| White Other | 0.54 (0.5 - 0.58) | 0.86 (0.79 - 0.93) | 0.42 (0.39 - 0.46) | 0.72 (0.71 - 0.74) | 0.81 (0.77 - 0.86) | 0.9 (0.85 - 0.95) | 1.04 (0.96 - 1.13) | 0.93 (0.91 - 0.95) | 0.81 (0.76 - 0.86) | 0.61 (0.56 - 0.67) | 0.74 (0.72 - 0.76) | 1.05 (0.96 - 1.16) |
| Latin American | 0.33 (0.27 - 0.4) | 0.5 (0.39 - 0.63) | 0.16 (0.12 - 0.22) | 0.77 (0.73 - 0.81) | 0.9 (0.8 - 1.01) | 0.75 (0.66 - 0.85) | 0.82 (0.68 - 1) | 1.13 (1.09 - 1.17) | 0.75 (0.65 - 0.87) | 0.22 (0.16 - 0.3) | 0.95 (0.91 - 1) | 1.12 (0.88 - 1.42) |
| **Smoking status** | | | | | | | | | | | | |
| Never smoked | - | - | - | - | - | - | - | - | - | - | - | - |
| Ever smoked | 4.34 (4.05 - 4.65) | 0.99 (0.94 - 1.05) | 2.25 (2.1 - 2.4) | 1.47 (1.44 - 1.5) | 1.35 (1.3 - 1.41) | 1.25 (1.2 - 1.3) | 1.52 (1.43 - 1.62) | 1.11 (1.09 - 1.12) | 1.09 (1.05 - 1.13) | 10.78 (9.52 - 12.21) | 1.48 (1.46 - 1.51) | 0.98 (0.92 - 1.04) |
| **Index of relative deprivation** | | | | | | | | | | | | |
| Most deprived | - | - | - | - | - | - | - | - | - | - | - | - |
| 2nd | 0.86 (0.8 - 0.93) | 1 (0.91 - 1.09) | 1.14 (1 - 1.29) | 0.95 (0.9 - 1) | 0.98 (0.92 - 1.04) | 1 (0.95 - 1.06) | 0.98 (0.9 - 1.06) | 1.01 (0.97 - 1.04) | 0.93 (0.87 - 0.99) | 0.79 (0.73 - 0.85) | 0.91 (0.86 - 0.96) | 1.06 (0.9 - 1.24) |
| 3rd | 0.77 (0.7 - 0.84) | 1.01 (0.92 - 1.12) | 1.26 (1.11 - 1.43) | 0.95 (0.9 - 1) | 0.93 (0.86 - 1) | 1.03 (0.97 - 1.1) | 0.9 (0.82 - 0.98) | 0.99 (0.96 - 1.03) | 0.9 (0.84 - 0.96) | 0.63 (0.58 - 0.7) | 0.85 (0.81 - 0.9) | 1.07 (0.9 - 1.28) |
| 4th | 0.73 (0.64 - 0.84) | 0.99 (0.86 - 1.14) | 1.24 (1.09 - 1.41) | 0.95 (0.89 - 1.02) | 0.97 (0.88 - 1.08) | 1.02 (0.93 - 1.11) | 0.84 (0.73 - 0.96) | 1 (0.97 - 1.04) | 0.87 (0.78 - 0.96) | 0.46 (0.39 - 0.54) | 0.81 (0.75 - 0.87) | 1.05 (0.83 - 1.33) |
| 5th | 0.68 (0.52 - 0.89) | 1.08 (0.81 - 1.45) | 1.25 (1.1 - 1.42) | 0.91 (0.82 - 1.01) | 1.13 (0.94 - 1.35) | 0.94 (0.78 - 1.14) | 0.79 (0.58 - 1.08) | 1.02 (0.98 - 1.06) | 0.92 (0.75 - 1.14) | 0.5 (0.35 - 0.71) | 0.76 (0.68 - 0.85) | 1.09 (0.71 - 1.69) |
| **Year joining cohort** | | | | | | | | | | | | |
| Before 2006 | - | - | - | - | - | - | - | - | - | - | - | - |
| 2006-2011 | 1 (0.94 - 1.07) | 0.72 (0.65 - 0.79) | 1.06 (0.99 - 1.14) | 0.96 (0.93 - 0.98) | 0.79 (0.75 - 0.84) | 0.79 (0.74 - 0.83) | 0.8 (0.73 - 0.87) | 0.85 (0.84 - 0.87) | 0.58 (0.54 - 0.62) | 0.76 (0.69 - 0.84) | 0.82 (0.8 - 0.84) | 0.84 (0.75 - 0.94) |
| 2011-2016 | 0.78 (0.72 - 0.84) | 0.75 (0.67 - 0.84) | 0.48 (0.44 - 0.54) | 1.16 (1.13 - 1.19) | 0.74 (0.7 - 0.79) | 0.71 (0.66 - 0.76) | 0.74 (0.66 - 0.82) | 0.76 (0.75 - 0.78) | 0.54 (0.5 - 0.59) | 0.67 (0.59 - 0.76) | 0.73 (0.72 - 0.75) | 0.75 (0.65 - 0.86) |
| 2016-2021 | 0.59 (0.52 - 0.66) | 0.59 (0.5 - 0.71) | 0.55 (0.48 - 0.62) | 1.18 (1.14 - 1.21) | 0.78 (0.72 - 0.85) | 0.6 (0.54 - 0.67) | 0.66 (0.56 - 0.78) | 0.64 (0.63 - 0.66) | 0.5 (0.44 - 0.56) | 0.42 (0.34 - 0.54) | 0.59 (0.57 - 0.61) | 0.52 (0.41 - 0.65) |
| 2021-2022 | 0.22 (0.11 - 0.42) | 0.35 (0.14 - 0.84) | 0.33 (0.2 - 0.57) | 0.38 (0.32 - 0.44) | 0.19 (0.12 - 0.31) | 0.05 (0.01 - 0.19) | 0.25 (0.09 - 0.66) | 0.22 (0.18 - 0.26) | 0.17 (0.08 - 0.37) | 0.2 (0.05 - 0.79) | 0.08 (0.05 - 0.11) | 0.07 (0.01 - 0.5) |

| Variable | Depression | Diabetes | Epilepsy | Heart Failure | HIV/AIDS | Hypertension | Inflammatory Bowel Disease | Learning Disabilities | Liver Disease | Lupus | Serious Mental Illness | Moderate obesity | Morbid Obesity |
|---|---|---|---|---|---|---|---|---|---|---|---|---|---|
| Total events | 30985 | 16929 | 1439 | 4379 | 1913 | 27522 | 1789 | 810 | 2036 | 286 | 3429 | 47201 | 10252 |
| **Sex** | | | | | | | | | | | | | |
| Male | - | - | - | - | - | - | - | - | - | - | - | - | - |
| Female | 1.66 (1.62 - 1.7) | 0.96 (0.93 - 0.99) | 0.83 (0.74 - 0.92) | 0.87 (0.82 - 0.92) | 0.23 (0.21 - 0.26) | 1.01 (0.99 - 1.03) | 1.04 (0.95 - 1.15) | 0.71 (0.62 - 0.82) | 0.71 (0.65 - 0.78) | 6.96 (4.92 - 9.86) | 0.9 (0.84 - 0.96) | 1.69 (1.66 - 1.73) | 2.43 (2.32 - 2.53) |
| **Ethnic group** | | | | | | | | | | | | | |
| White British | - | - | - | - | - | - | - | - | - | - | - | - | - |
| Black/African/Caribbean/Black British | 0.8 (0.77 - 0.83) | 2.65 (2.54 - 2.76) | 1 (0.88 - 1.15) | 1.16 (1.07 - 1.25) | 1.49 (1.32 - 1.69) | 2.05 (1.99 - 2.12) | 0.57 (0.49 - 0.65) | 0.81 (0.68 - 0.97) | 0.88 (0.78 - 0.99) | 3.48 (2.49 - 4.87) | 1.9 (1.74 - 2.07) | 1.89 (1.84 - 1.94) | 1.75 (1.66 - 1.84) |
| Asian/Asian British | 0.74 (0.71 - 0.78) | 3.67 (3.48 - 3.88) | 0.58 (0.45 - 0.75) | 1.23 (1.1 - 1.39) | 0.45 (0.34 - 0.6) | 1.55 (1.48 - 1.63) | 0.77 (0.64 - 0.94) | 0.51 (0.37 - 0.71) | 1.08 (0.91 - 1.28) | 3.49 (2.24 - 5.44) | 1.08 (0.93 - 1.26) | 0.97 (0.93 - 1.01) | 0.68 (0.61 - 0.75) |
| Mixed/Multiple ethnic group | 1.13 (1.07 - 1.18) | 1.98 (1.82 - 2.16) | 1.04 (0.81 - 1.34) | 1.19 (1 - 1.42) | 1.41 (1.13 - 1.76) | 1.49 (1.39 - 1.59) | 0.58 (0.44 - 0.76) | 1 (0.75 - 1.34) | 0.8 (0.62 - 1.04) | 3.03 (1.79 - 5.12) | 1.91 (1.66 - 2.18) | 1.39 (1.33 - 1.45) | 1.38 (1.26 - 1.52) |
| Other ethnic group | 0.78 (0.72 - 0.85) | 2.04 (1.85 - 2.26) | 0.67 (0.45 - 1) | 0.99 (0.77 - 1.28) | 0.87 (0.63 - 1.21) | 1.17 (1.07 - 1.27) | 0.64 (0.45 - 0.91) | 0.47 (0.27 - 0.84) | 0.72 (0.51 - 1.01) | 1.27 (0.46 - 3.52) | 1.08 (0.86 - 1.37) | 1.1 (1.03 - 1.17) | 0.99 (0.86 - 1.15) |
| White Other | 0.68 (0.66 - 0.7) | 1.16 (1.1 - 1.22) | 0.68 (0.58 - 0.79) | 1.04 (0.95 - 1.15) | 1.16 (1.02 - 1.31) | 0.99 (0.95 - 1.03) | 0.78 (0.69 - 0.88) | 0.49 (0.39 - 0.62) | 0.91 (0.8 - 1.03) | 1.43 (0.95 - 2.14) | 0.73 (0.66 - 0.82) | 0.94 (0.91 - 0.97) | 0.82 (0.77 - 0.88) |
| Latin American | 0.76 (0.71 - 0.81) | 1.61 (1.47 - 1.77) | 0.73 (0.53 - 1.01) | 0.74 (0.58 - 0.95) | 2 (1.65 - 2.43) | 0.93 (0.86 - 1) | 0.62 (0.46 - 0.83) | 0.36 (0.21 - 0.62) | 0.65 (0.49 - 0.87) | 2.28 (1.18 - 4.38) | 0.92 (0.74 - 1.14) | 1.42 (1.36 - 1.49) | 0.89 (0.79 - 1) |
| **Smoking status** | | | | | | | | | | | | | |
| Never smoked | - | - | - | - | - | - | - | - | - | - | - | - | - |
| Ever smoked | 1.84 (1.79 - 1.88) | 1.34 (1.3 - 1.39) | 1.38 (1.24 - 1.55) | 1.25 (1.17 - 1.33) | 1.28 (1.17 - 1.41) | 1.07 (1.04 - 1.09) | 1.32 (1.19 - 1.45) | 0.65 (0.56 - 0.75) | 1.44 (1.31 - 1.58) | 1.7 (1.33 - 2.16) | 2.5 (2.32 - 2.7) | 1.1 (1.08 - 1.12) | 1.16 (1.11 - 1.21) |
| **Index of relative deprivation** | | | | | | | | | | | | | |
| Most deprived | - | - | - | - | - | - | - | - | - | - | - | - | - |
| 2nd | 0.91 (0.86 - 0.95) | 0.89 (0.84 - 0.94) | 0.77 (0.66 - 0.9) | 0.99 (0.9 - 1.1) | 0.93 (0.8 - 1.08) | 0.99 (0.94 - 1.04) | 0.96 (0.84 - 1.09) | 0.7 (0.57 - 0.87) | 0.9 (0.79 - 1.02) | 0.84 (0.63 - 1.11) | 0.84 (0.75 - 0.94) | 0.95 (0.89 - 1.01) | 0.82 (0.76 - 0.87) |
| 3rd | 0.84 (0.8 - 0.89) | 0.75 (0.71 - 0.8) | 0.75 (0.63 - 0.9) | 0.92 (0.82 - 1.02) | 0.81 (0.68 - 0.97) | 0.9 (0.86 - 0.95) | 0.99 (0.86 - 1.14) | 0.51 (0.39 - 0.66) | 0.76 (0.66 - 0.88) | 0.64 (0.45 - 0.92) | 0.7 (0.61 - 0.8) | 0.84 (0.78 - 0.9) | 0.68 (0.63 - 0.74) |
| 4th | 0.78 (0.73 - 0.85) | 0.72 (0.65 - 0.79) | 0.65 (0.5 - 0.86) | 0.78 (0.66 - 0.92) | 0.72 (0.54 - 0.95) | 0.87 (0.83 - 0.92) | 1.07 (0.87 - 1.31) | 0.41 (0.27 - 0.64) | 0.67 (0.53 - 0.84) | 0.96 (0.57 - 1.6) | 0.61 (0.49 - 0.74) | 0.82 (0.77 - 0.87) | 0.53 (0.47 - 0.6) |
| 5th | 0.77 (0.68 - 0.87) | 0.7 (0.58 - 0.85) | 0.52 (0.28 - 0.97) | 0.6 (0.4 - 0.91) | 0.44 (0.23 - 0.82) | 0.89 (0.84 - 0.93) | 0.8 (0.52 - 1.23) | 0.24 (0.08 - 0.79) | 0.68 (0.42 - 1.1) | 0.89 (0.28 - 2.86) | 0.49 (0.31 - 0.76) | 0.74 (0.7 - 0.79) | 0.54 (0.42 - 0.69) |
| **Year joining cohort** | | | | | | | | | | | | | |
| Before 2006 | - | - | - | - | - | - | - | - | - | - | - | - | - |
| 2006-2011 | 0.85 (0.82 - 0.87) | 0.78 (0.75 - 0.82) | 0.84 (0.73 - 0.97) | 0.69 (0.62 - 0.77) | 1.27 (1.13 - 1.42) | 0.76 (0.73 - 0.79) | 0.94 (0.82 - 1.06) | 0.63 (0.52 - 0.76) | 0.93 (0.83 - 1.05) | 0.61 (0.44 - 0.85) | 0.96 (0.88 - 1.06) | 0.78 (0.76 - 0.8) | 0.76 (0.72 - 0.8) |
| 2011-2016 | 0.87 (0.84 - 0.9) | 0.77 (0.73 - 0.81) | 0.72 (0.61 - 0.86) | 0.72 (0.63 - 0.81) | 0.92 (0.81 - 1.05) | 0.68 (0.65 - 0.71) | 0.85 (0.74 - 0.98) | 0.37 (0.29 - 0.47) | 0.96 (0.84 - 1.09) | 0.57 (0.39 - 0.84) | 0.86 (0.77 - 0.95) | 0.66 (0.64 - 0.68) | 0.67 (0.63 - 0.71) |
| 2016-2021 | 0.98 (0.95 - 1.02) | 0.57 (0.53 - 0.63) | 0.73 (0.59 - 0.91) | 0.67 (0.55 - 0.8) | 0.62 (0.5 - 0.76) | 0.58 (0.54 - 0.62) | 0.93 (0.79 - 1.11) | 0.4 (0.31 - 0.53) | 0.61 (0.49 - 0.77) | 0.42 (0.23 - 0.74) | 0.96 (0.84 - 1.09) | 0.58 (0.56 - 0.61) | 0.54 (0.49 - 0.59) |
| 2021-2022 | 0.44 (0.37 - 0.52) | 0.1 (0.05 - 0.2) | 0.16 (0.04 - 0.63) | 0.17 (0.04 - 0.67) | 0.22 (0.07 - 0.7) | 0.16 (0.1 - 0.25) | 0.37 (0.16 - 0.83) | 0 (0 - Inf) | 0.1 (0.01 - 0.71) | 0.39 (0.05 - 2.85) | 0.36 (0.19 - 0.67) | 0.21 (0.16 - 0.28) | 0.18 (0.1 - 0.3) |

| Variable | Multiple Sclerosis | Osteoporosis | Osteoarthritis | Peripheral Vascular Disease | Parkinson's | Rheumatoid Arthritis | Substance Dependency | Substance Use Excluding Dependency | Viral Hepatitis (B & C) |
|---|---|---|---|---|---|---|---|---|---|
| Total events | 340 | 2634 | 17711 | | | | 3479 | 5671 | 2444 |
| **Sex** | | | | | | | | | |
| Male | - | - | - | - | - | - | - | - | - |
| Female | 2.04 (1.63 - 2.56) | 4.82 (4.36 - 5.34) | 2.04 (1.98 - 2.1) | 0.64 (0.58 — 0.71) | 0.61 (0.52 — 0.72) | 2.71 (2.42 — 3.03) | 0.5 (0.46 - 0.53) | 0.71 (0.68 - 0.75) | 0.61 (0.56 - 0.67) |
| **Ethnic group** | | | | | | | | | |
| White British | - | - | - | - | - | - | - | - | - |
| Black/African/Caribbean/Black British | 0.76 (0.56 - 1.01) | 0.27 (0.23 - 0.31) | 1.31 (1.26 - 1.37) | 0.76 (0.67 — 0.86) | 0.69 (0.56 — 0.85) | 1.17 (1.02 — 1.34) | 0.86 (0.78 - 0.94) | 0.96 (0.9 - 1.03) | 2.39 (2.14 - 2.67) |
| Asian/Asian British | 0.47 (0.26 - 0.82) | 0.78 (0.66 - 0.91) | 1.27 (1.19 - 1.34) | 0.92 (0.76 — 1.11) | 1.1 (0.83 — 1.45) | 1.52 (1.25 — 1.83) | 0.39 (0.31 - 0.48) | 0.52 (0.45 - 0.6) | 2.38 (2.04 - 2.77) |
| Mixed/Multiple ethnic group | 0.85 (0.51 - 1.43) | 0.45 (0.33 - 0.61) | 1.11 (1.02 - 1.21) | 0.8 (0.6 — 1.07) | 0.96 (0.59 — 1.54) | 0.97 (0.72 — 1.31) | 1.17 (1.01 - 1.34) | 1.25 (1.13 - 1.39) | 1.78 (1.45 - 2.18) |
| Other ethnic group | 0.82 (0.4 - 1.66) | 0.69 (0.49 - 0.97) | 1.07 (0.96 - 1.19) | 0.48 (0.29 — 0.77) | 0.65 (0.31 — 1.38) | 1.33 (0.95 — 1.86) | 0.67 (0.53 - 0.85) | 0.73 (0.61 - 0.87) | 1.47 (1.11 - 1.94) |
| White Other | 0.71 (0.53 - 0.95) | 0.85 (0.76 - 0.95) | 0.94 (0.9 - 0.99) | 0.89 (0.78 — 1.02) | 1.03 (0.82 — 1.29) | 1.02 (0.88 — 1.19) | 0.65 (0.59 - 0.71) | 0.57 (0.53 - 0.62) | 1.44 (1.28 - 1.63) |
| Latin American | 0.39 (0.17 - 0.89) | 0.65 (0.47 - 0.88) | 1.22 (1.12 - 1.33) | 0.63 (0.43 — 0.91) | 0.36 (0.15 — 0.86) | 1.69 (1.32 — 2.17) | 0.55 (0.44 - 0.69) | 0.67 (0.57 - 0.78) | 0.84 (0.63 - 1.11) |
| **Smoking status** | | | | | | | | | |
| Never smoked | - | - | - | - | - | - | - | - | - |
| Ever smoked | 1.62 (1.29 - 2.03) | 0.93 (0.85 - 1.01) | 1.06 (1.03 - 1.1) | 2.99 (2.64 — 3.38) | 0.75 (0.64 — 0.88) | 1.49 (1.34 — 1.67) | 7.01 (6.31 - 7.79) | 4.48 (4.19 - 4.8) | 1.42 (1.31 - 1.54) |
| **Index of relative deprivation** | | | | | | | | | |
| Most deprived | - | - | - | - | - | - | - | - | - |
| 2nd | 0.78 (0.59 - 1.04) | 1.17 (1.02 - 1.34) | 1.02 (0.97 - 1.07) | 1.05 (0.91 — 1.21) | 0.9 (0.69 — 1.19) | 1.07 (0.92 — 1.24) | 0.9 (0.8 - 1.02) | 0.85 (0.76 - 0.95) | 0.89 (0.78 - 1.02) |
| 3rd | 0.77 (0.56 - 1.06) | 1.19 (1.03 - 1.39) | 0.96 (0.91 - 1.01) | 0.92 (0.79 — 1.08) | 0.97 (0.73 — 1.28) | 0.87 (0.73 — 1.02) | 0.81 (0.71 - 0.92) | 0.8 (0.72 - 0.89) | 0.84 (0.73 - 0.97) |
| 4th | 0.94 (0.6 - 1.48) | 1.44 (1.17 - 1.76) | 0.92 (0.85 - 1) | 0.71 (0.6 — 0.84) | 1.02 (0.77 — 1.35) | 0.98 (0.84 — 1.15) | 0.75 (0.65 - 0.85) | 0.8 (0.71 - 0.89) | 0.76 (0.65 - 0.88) |
| 5th | 0.99 (0.43 - 2.29) | 1.76 (1.21 - 2.56) | 0.94 (0.8 - 1.12) | 0.74 (0.63 — 0.86) | 1.09 (0.84 — 1.43) | 0.9 (0.77 — 1.06) | 0.68 (0.59 - 0.78) | 0.65 (0.58 - 0.73) | 0.61 (0.52 - 0.72) |
| **Year joining cohort** | | | | | | | | | |
| Before 2006 | - | - | - | - | - | - | - | - | - |
| 2006-2011 | 0.8 (0.6 - 1.07) | 0.78 (0.67 - 0.91) | 0.74 (0.71 - 0.78) | 0.73 (0.62 — 0.86) | 0.81 (0.62 — 1.06) | 0.82 (0.7 — 0.95) | 0.89 (0.82 - 0.98) | 0.91 (0.85 - 0.98) | 1.31 (1.19 - 1.45) |
| 2011-2016 | 0.78 (0.57 - 1.06) | 0.79 (0.66 - 0.95) | 0.72 (0.68 - 0.76) | 0.69 (0.56 — 0.84) | 0.65 (0.46 — 0.92) | 0.85 (0.72 — 1.01) | 0.83 (0.75 - 0.91) | 0.98 (0.9 - 1.05) | 1 (0.88 - 1.12) |
| 2016-2021 | 0.66 (0.43 - 1.01) | 0.59 (0.44 - 0.79) | 0.52 (0.48 - 0.58) | 0.62 (0.45 — 0.86) | 0.34 (0.17 — 0.68) | 0.77 (0.6 — 0.98) | 0.68 (0.59 - 0.78) | 0.93 (0.85 - 1.03) | 0.6 (0.49 - 0.73) |
| 2021-2022 | 0.32 (0.04 - 2.28) | 0 (0 - Inf) | 0.16 (0.08 - 0.29) | 0 (0 — Inf) | 0 (0 — Inf) | 0 (0 — Inf) | 0.36 (0.19 - 0.69) | 0.35 (0.22 - 0.58) | 0.14 (0.03 - 0.55) |

Table S4: Sensitivity analysis of the effect of imputing missing ethnic group data as White British or Latin American. n=100,000, randomly sampled from the full population.

| | Multivariable hazard ratios for Latin American population | | |
|---|---|---|---|
| **Long-term condition** | **Sample of 100,000 complete case analysis** | **Missing ethnicity data imputed as White British** | **Missing ethnicity data imputed as Latin American** |
| **Alcohol Dependence** | 0.31 (0.17 — 0.56) | 0.66 (0.54 — 0.81) | 0.45 (0.37 — 0.53) |
| **Atrial Fibrillation** | 0.75 (0.43 — 1.31) | 0.9 (0.7 — 1.15) | 0.47 (0.38 — 0.58) |
| **Anxiety Disorders** | 0.73 (0.63 — 0.85) | 0.86 (0.81 — 0.93) | 0.63 (0.59 — 0.67) |
| **Asthma** | 1.09 (0.79 — 1.5) | 1.07 (0.91 — 1.27) | 0.72 (0.62 — 0.84) |
| **Cancer** | 0.65 (0.44 — 0.95) | 1.11 (0.94 — 1.29) | 0.68 (0.6 — 0.79) |
| **Coronary Heart Disease** | 1.08 (0.65 — 1.8) | 1.29 (1.02 — 1.65) | 0.71 (0.58 — 0.89) |
| **Chronic Kidney Disease 3-5** | 1.1 (0.77 — 1.57) | 0.96 (0.79 — 1.15) | 0.55 (0.47 — 0.65) |
| **COPD** | 0.26 (0.11 — 0.63) | 0.7 (0.54 — 0.92) | 0.39 (0.31 — 0.49) |
| **Chronic Pain** | 1.1 (0.96 — 1.26) | 0.89 (0.82 — 0.96) | 0.57 (0.53 — 0.61) |
| **Dementia** | 1.3 (0.71 — 2.35) | 1.45 (1.1 — 1.91) | 0.65 (0.51 — 0.83) |
| **Depression** | 0.89 (0.74 — 1.06) | 0.8 (0.73 — 0.88) | 0.56 (0.52 — 0.61) |
| **Diabetes** | 1.63 (1.23 — 2.16) | 1.49 (1.28 — 1.75) | 0.84 (0.73 — 0.97) |
| **Epilepsy** | 0.6 (0.22 — 1.65) | 1.24 (0.83 — 1.85) | 0.63 (0.44 — 0.91) |
| **Heart Failure** | 0.91 (0.48 — 1.72) | 1.13 (0.85 — 1.5) | 0.68 (0.53 — 0.86) |
| **HIV/AIDS** | 3.1 (1.72 — 5.59) | 1.93 (1.33 — 2.8) | 1.28 (0.88 — 1.84) |
| **Inflammatory Bowel Disease** | 0.59 (0.21 — 1.62) | 0.83 (0.55 — 1.24) | 0.66 (0.46 — 0.94) |
| **Learning Disabilities** | 0.6 (0.18 — 1.96) | 0.58 (0.29 — 1.15) | 0.34 (0.2 — 0.58) |
| **Liver Disease** | 1.26 (0.65 — 2.46) | 1.3 (0.92 — 1.85) | 0.86 (0.63 — 1.19) |
| **Lupus** | 1.52 (0.18 — 12.51) | 1.65 (0.58 — 4.72) | 1.2 (0.43 — 3.34) |
| **Serious Mental Illness** | 1.21 (0.7 — 2.12) | 0.79 (0.58 — 1.09) | 0.61 (0.47 — 0.81) |
| **Morbid Obesity** | 0.81 (0.55 — 1.18) | 1 (0.84 — 1.21) | 0.68 (0.57 — 0.8) |
| **Multiple Sclerosis** | 0.76 (0.1 — 5.87) | 0.75 (0.29 — 1.9) | 0.79 (0.36 — 1.76) |
| **Osteoporosis** | 0.71 (0.33 — 1.54) | 1 (0.72 — 1.39) | 0.42 (0.31 — 0.56) |
| **Osteoarthritis** | 1.42 (1.12 — 1.8) | 1.18 (1.03 — 1.36) | 0.65 (0.57 — 0.74) |
| **Peripheral Vascular Disease** | 0.5 (0.16 — 1.6) | 1.07 (0.73 — 1.56) | 0.75 (0.55 — 1.03) |
| **Parkinson's** | 0 (0 — Inf) | 1.02 (0.5 — 2.1) | 0.62 (0.35 — 1.09) |
| **Rheumatoid Arthritis** | 1.87 (0.91 — 3.81) | 1.05 (0.66 — 1.66) | 0.79 (0.53 — 1.19) |

| | | | |
|---|---|---|---|
| **Sickle-Cell Disease** | 1.23 (0 — Inf) | 0 (0 — Inf) | 10741817941.43 (0 — Inf) |
| **Stroke** | 1.08 (0.61 — 1.9) | 1.07 (0.81 — 1.42) | 0.68 (0.54 — 0.87) |
| **Substance Dependency** | 0.45 (0.22 — 0.91) | 0.56 (0.41 — 0.75) | 0.4 (0.31 — 0.52) |
| **Transient Ischemic Attack** | 1.22 (0.53 — 2.82) | 1.15 (0.76 — 1.74) | 0.73 (0.51 — 1.04) |
| **Viral Hepatitis (B & C)** | 0.75 (0.3 — 1.89) | 1.48 (1.04 — 2.11) | 1.24 (0.88 — 1.74) |
| **Hypertension** | 1.08 (0.87 — 1.35) | 1.13 (1 — 1.27) | 0.59 (0.53 — 0.65) |
| **Substance Use** | 0.7 (0.46 — 1.06) | 0.59 (0.48 — 0.72) | 0.44 (0.37 — 0.53) |
| **Alcohol >14 units/week** | 0.26 (0.14 — 0.51) | 0.55 (0.43 — 0.7) | 0.27 (0.21 — 0.34) |
| **High Cholesterol** | 1.15 (1.03 — 1.27) | 1.26 (1.19 — 1.34) | 0.58 (0.56 — 0.62) |

Table S5: Cumulative incidence of multimorbidity by ethnic group, 2005-2022. Multimorbidity was defined as having had two or more long-term conditions (LTCs) diagnosed on separate days during follow up. Counted LTCs are: alcohol dependence; atrial fibrillation; anxiety disorders; asthma; cancer; coronary heart disease; chronic kidney disease stages 3-5; COPD; chronic pain; dementia; depression; diabetes; epilepsy; heart failure; HIV/AIDS; inflammatory bowel disease; learning disabilities; liver disease; lupus; serious mental illness; morbid obesity; multiple sclerosis; osteoporosis; osteoarthritis; peripheral vascular disease; Parkinson's disease; rheumatoid arthritis; sickle-cell disease; stroke; substance dependency; transient ischemic attack; viral hepatitis B and C.

| Ethnic group | n | Cumulative incidence of multimorbidity (%) |
|---|---|---|
| **Black/African/Caribbean/Black British** | 120067 | 26.7 |
| **White** | 287304 | 24.3 |
| **Mixed/Multiple ethnic group** | 31587 | 19.9 |
| **Asian/Asian British** | 56411 | 16.3 |
| **Other ethnic group** | 18654 | 14 |
| **White Other** | 185219 | 11.4 |
| **Latin American** | 28617 | 10.9 |
| **Missing ethnic group data** | 162579 | 9 |
